# Current State and Demographic Trends of Medically Underserved Populations in Rare Disease Research in the United States

**DOI:** 10.1101/2025.04.30.25326209

**Authors:** Lavanyaa Manjunatha, MS Saundarya, Deepika Dokuru, Nisha Venugopal, Jenifer Ngo Waldrop, Linda Goler Blount, Reena V Kartha, Harsha K Rajasimha

## Abstract

**Background:** Addressing inequities and health disparities for medically underserved populations (MUPs) is critical, as they already face systemic bias and barriers, such as historical mistrust of healthcare systems. To achieve health equity, we need systematic approaches to measuring, quantifying, and reporting diversity, equity, inclusion, and accessibility (DEIA) metrics.

**Methods:** The objective of the study was to analyze literature and clinical trials to summarize the current state of demographics and socioeconomic factors (SF) reporting for MUPs in US-based RD clinical research. PubMed, Cochrane Library, and ClinicalTrials.gov were searched (1983– 2023). A universal set of 30,303 unique RD terms from the Genetic and Rare Diseases Information Center, Orphanet, Rare-X, and ClinicalTrials.gov was used to filter publications and clinical trials. Publications that reported demographics or SFs, were US-based, and involved one or more RDs were included for analysis. Clinical trials that were US-based, involved an RD, and had study results posted were also included. Age, sex or gender, race, ethnicity, and SF data were extracted and analyzed using descriptive statistics. Race and ethnicity data were compared with the US census. The representation of MUPs in RD clinical research was assessed based on the frequency of publications and clinical trials reporting 13 variables.

**Results:** We reviewed 234 publications and 8475 RD clinical trials. Age was the most reported demographic variable (publications: 94%; clinical trials: 100%), followed by sex or gender (86.3%; 100%). Race (50%; 45.7%) and ethnicity (29.9%; 38.5%) were less frequently reported and often in a variable format in publications compared with the ClinicalTrials.gov database. At least one SF was reported in 15.8% of the publications and 0.2% of the trials. American Indian or Native Alaskan, Asian, Hispanic, and Latino participants were significantly underrepresented compared with the US census averages. Data were largely absent for other MUPs: lesbian, gay, bisexual, transgender, and queer or questioning individuals, rural residents, veterans, immigrants, and those affected by disability and poverty.

**Conclusions:** Significant gaps exist in demographics and SF reporting in RD clinical research, and several MUPs are underrepresented. Therefore, a framework to enhance DEIA in RD research is urgently needed.

## Background

Improving representation in clinical trials has been a major focus of United States (US) legislators and the Food and Drug Administration (FDA) in recent years (1–4). The FDA’s latest Diversity Action Plan (DAP) aims to enhance diversity in clinical trials, including in rare disease (RD) trials (4). Over 10,000 RDs collectively affect 30 million Americans (5,6), with a $997 billion economic burden (7), making them a public health concern. Marking the 40th anniversary of the Orphan Drug Act in 2023, the approval of >880 treatments for RDs in this period indicate progress yet highlight unmet needs (8,9). The FDA has several programs to accelerate RD research, including the Rare Disease Innovation Hub and the Support for Clinical Trials Advancing Rare Disease Therapeutics Pilot Program (10–14). For these programs to be effective, accurate assessment and tracking of the current status of diversity, equity, inclusion, and accessibility (DEIA, Box 1) in US-based RD research is necessary.

The FDA’s Drug Trials Snapshot (DTS, 2015–2019) revealed underrepresentation in global RD trials, with 11% Asian, 9% Black or African American, 1% American Indian or Alaska Native, 6% Hispanic or Latino, and 34% missing ethnicity data (15). In cystic fibrosis (CF) trials, Black or African American, Asian, and Hispanic or Latino participants were included at lower rates than in the US CF Foundation registry (16). Similar underrepresentation has been observed in US trials of muscular dystrophy, hemophilia, and acute leukemia (17–19), indicating underrepresentation is prevalent in RD research.

This persistent underrepresentation could be explained by common RD features: low prevalence, varying prevalence in medically underserved populations (MUPs), complex natural history, dispersed geographic patient distribution, diagnostic difficulties, patient registry reliance, and lack of awareness among care providers and patients (20). Given the small trial sizes of RDs (21), defining what DEIA entails in these contexts is vital. Although the US census serves as a representation benchmark (22,23), trial diversity should also be assessed based on prevalence and epidemiology (24). Using estimates in the absence of epidemiology data (25) adds another layer of complexity to ensure the inclusion of all affected patients in RDs.

Demographic and socioeconomic factors (SFs) disparities are evident in RD research. Only 20% of RD trials had child participants (26), despite 50–70% of RDs affecting them (27,28). Female patients with RDs, often dismissed by healthcare providers (29), face delayed diagnosis and lower health-related quality of life than male patients (30,31). Disparities in diagnostic delays and greater disease severity are apparent for Black or African American patients with amyotrophic lateral sclerosis (32). Hispanic patients with CF had lower pulmonary function than non-Hispanic patients with CF (33). Further, SFs impact the mortality and survival of patients with pulmonary arterial hypertension (34). Delayed diagnosis of RDs results in direct and indirect costs of $517K per patient (35). These data underscore the urgent need to enhance DEIA in RDs, emphasizing the moral, economic, and therapeutic imperatives for diverse representation.

Understanding the current landscape of MUP participation is essential to advancing DEIA in RD research. Except for the FDA’s DTS report, which provides some data about representation, no other studies have reported such data (36). This study aimed to quantify the reporting of demographics and SFs in RD research to enhance DEIA for MUPs through evidence-based frameworks. We report the findings from a detailed literature assessment (2010–2023) and an analysis of US-based clinical trials (1983–2023) registered on ClinicalTrials.gov.

### Box 1

**Definitions**

(a) ***diversity^a^*** means the practice of including the many communities, identities, races, ethnicities, backgrounds, abilities, cultures, and beliefs of the American people, including underserved communities.
(b) ***equity^a,b^*** means the consistent and systematic fair, just, and impartial treatment of all individuals, including individuals who belong to underserved communities that have been denied such treatment.
(c) ***inclusion^a^*** means the recognition, appreciation, and use of the talents and skills of employees of all backgrounds.
(d) ***accessibility*^a^** means the design, construction, development, and maintenance of facilities, information and communication technology, programs, and services so that all people, including people with disabilities, can fully and independently use them. Accessibility includes the provision of accommodations and modifications to ensure equal access to employment and participation in activities for people with disabilities, the reduction or elimination of physical and attitudinal barriers to equitable opportunities, a commitment to ensuring that people with disabilities can independently access every outward-facing and internal activity or electronic space, and the pursuit of best practices such as universal design.
(e) ***medically underserved populations (MUPs)^c^*** are populations facing a shortage of primary healthcare services for a specific population subset within a geographic area. These groups may face economic, cultural, or language barriers to healthcare. MUPs could include people experiencing homelessness, low-income people, people eligible for Medicaid, Native Americans, and migrant farm workers.
(d) ***medically underserved areas (MUAs)*** have a shortage of primary health services within geographic areas such as a whole county, a group of neighboring counties, a group of urban census tracts, a group of county or civil divisions
(f) ***underserved communities^a^***refer to populations sharing a particular characteristic, as well as geographic communities, who have been systematically denied a full opportunity to participate in aspects of economic, social, and civic life. In the context of the Federal workforce, this term includes individuals who belong to communities of color, such as Black and African American, Hispanic and Latino, Native American, Alaska Native and Indigenous, Asian American, Native Hawaiian and Pacific Islander, Middle Eastern, and North African persons. It also includes individuals who belong to communities that face discrimination based on sex, sexual orientation, and gender identity (including lesbian, gay, bisexual, transgender, queer, gender non-conforming, and non-binary (LGBTQ+) persons); persons who face discrimination based on pregnancy or pregnancy-related conditions; parents; and caregivers. It also includes individuals who belong to communities that face discrimination based on their religion or disability; first-generation professionals or first-generation college students; individuals with limited English proficiency; immigrants; individuals who belong to communities that may face employment barriers based on older age or former incarceration; persons who live in rural areas; veterans and military spouses; and persons otherwise adversely affected by persistent poverty, discrimination, or inequality. Individuals may belong to more than one underserved community and face intersecting barriers. ^a^ Executive order 14,035 on diversity, equity, inclusion, and accessibility in the federal workforce. ^b^ Executive order 13,985 on advancing racial equity and support for underserved communities through the federal government. ^c^Health Resource and Services Administration

## Methods

The study assessed 13 demographic and SF variables in RD research by reviewing the literature and analyzing the ClinicalTrials.gov database; methods are summarized below and detailed in the Supplementary Information. This study followed the Standards for Quality Improvement Reporting Excellence and Strengthening the Reporting of Observational Studies in Epidemiology guidelines (37,38).

### Literature assessment

PubMed and the Cochrane Library were searched using specific terms (Additional file 1: Table S1). Screening and filtering details are in Additional file 1: Supplementary Methods 2, Figure S1A, and Table S2. Publications were included if they reported demographics or SFs, involved one or more RD conditions, and were conducted in the US. Qualitative and quantitative data were extracted from publications satisfying our defined inclusion criteria.

### ClinicalTrials.gov database extraction and analysis

All clinical trials data (8 December 2023) and study information for US-based trials were downloaded from ClinicalTrials.gov (Filters: Location=USA, Study results=With Study results, Date range=01/01/1983–08/12/2023) (39). RD trials were identified by matching the “condition” section of clinical trials with a comprehensive list of RD terms generated from the Genetic and Rare Diseases Information Center (GARD), Orphanet, Rare-X, and the ClinicalTrials.gov classical website (find studies by topic/RDs) (40–43). See Supplementary Information for details. Trials with SFs were identified using keywords (Additional file 1: Table S3).

### Study Outcomes

The primary outcomes were representation of MUPs (historically underserved racial and ethnic groups, multiracial groups, lesbian, gay, bisexual, transgender, queer or questioning (LGBTQ+) individuals, caregivers, rural residents, veterans, religious minorities, immigrants, people living with disability, and those affected by poverty). We also gathered the frequency of reporting in publications and clinical trials of the variables: age, sex, gender, sexual orientation, race, ethnicity, and SFs (language, education, employment, income, residence location, health insurance, relationship status, social support, and disability allowance).

### Statistical Analysis

Descriptive statistics were used to summarize the publication or trial characteristics, demographics, and SFs. Racial and ethnic proportions from trials reporting them in the National Institute of Health (NIH)/Office of Management and Budget (OMB) format ((44) were compared with the weighted average percentages of US census data (1990–2022). The weights were based on trial participant distribution in the census bins. These weighted average percentages are referred to as US census averages from this point forward.

Chi-squared tests assessed significant differences in race and ethnicity distributions between the trials and the census. Bonferroni correction was used to adjust the significance threshold for multiple comparisons in the post hoc analyses. All statistical analyses were performed using Python 3.

## Results

### Literature assessment

Of the 16,187 selected publications, 1279 underwent full-text reviews to assess inclusion criteria. The analysis included 234 publications (56% US-only; 44% multinational; Additional file 1: Figure S1A). Table 1 presents the publication characteristics, participant type, demographics, and SFs reported in RD publications.

**Table 1:**
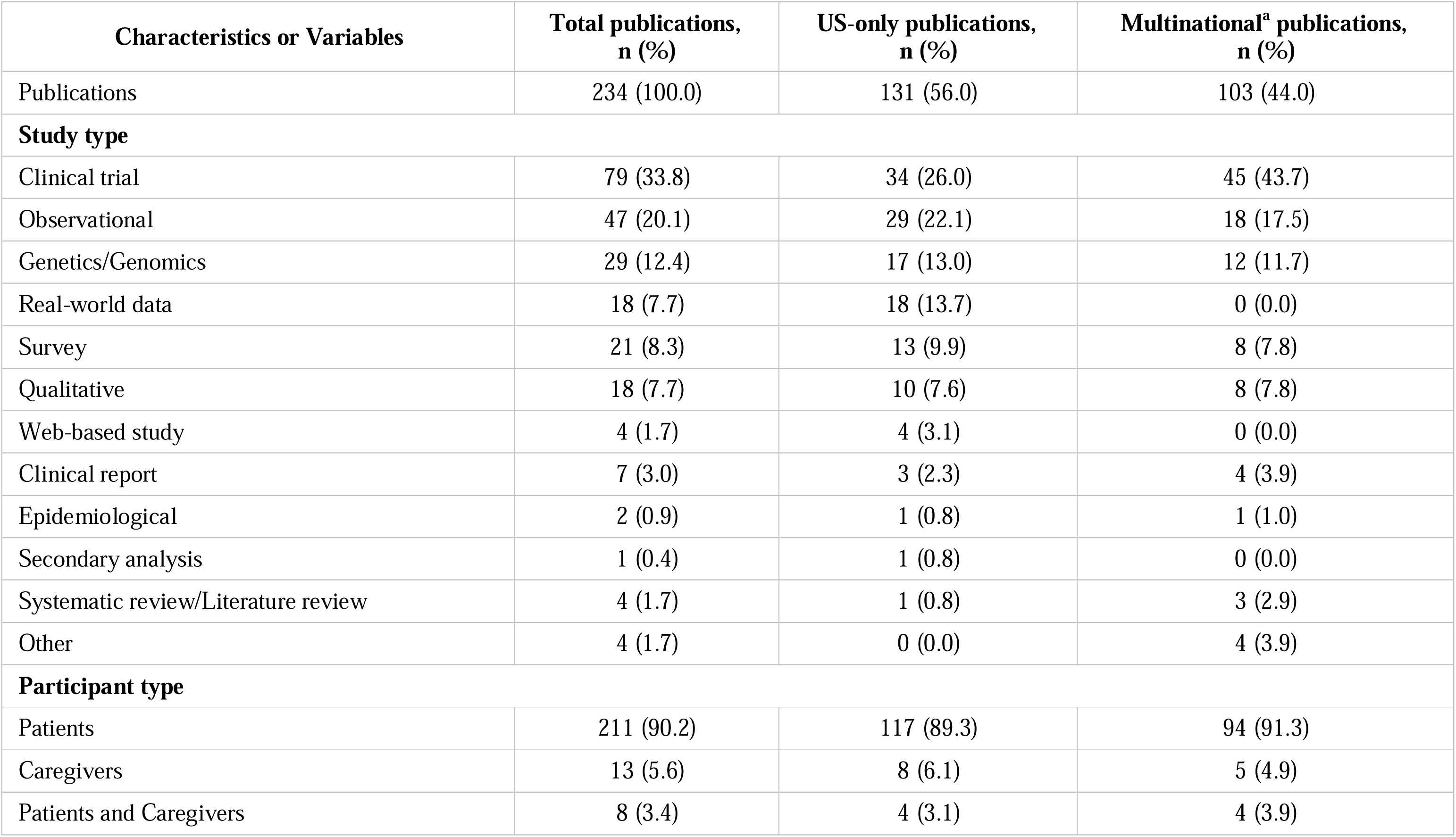
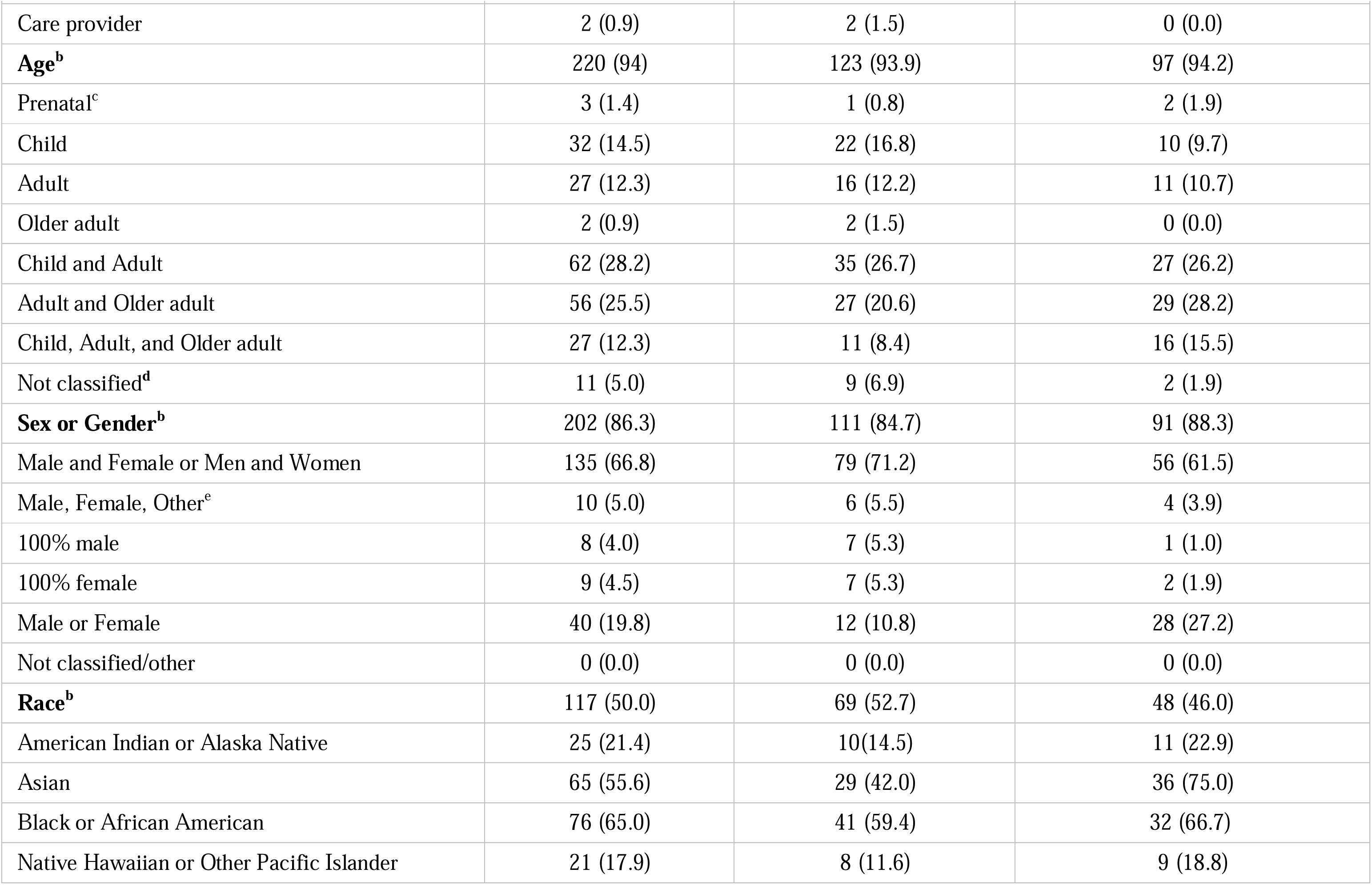
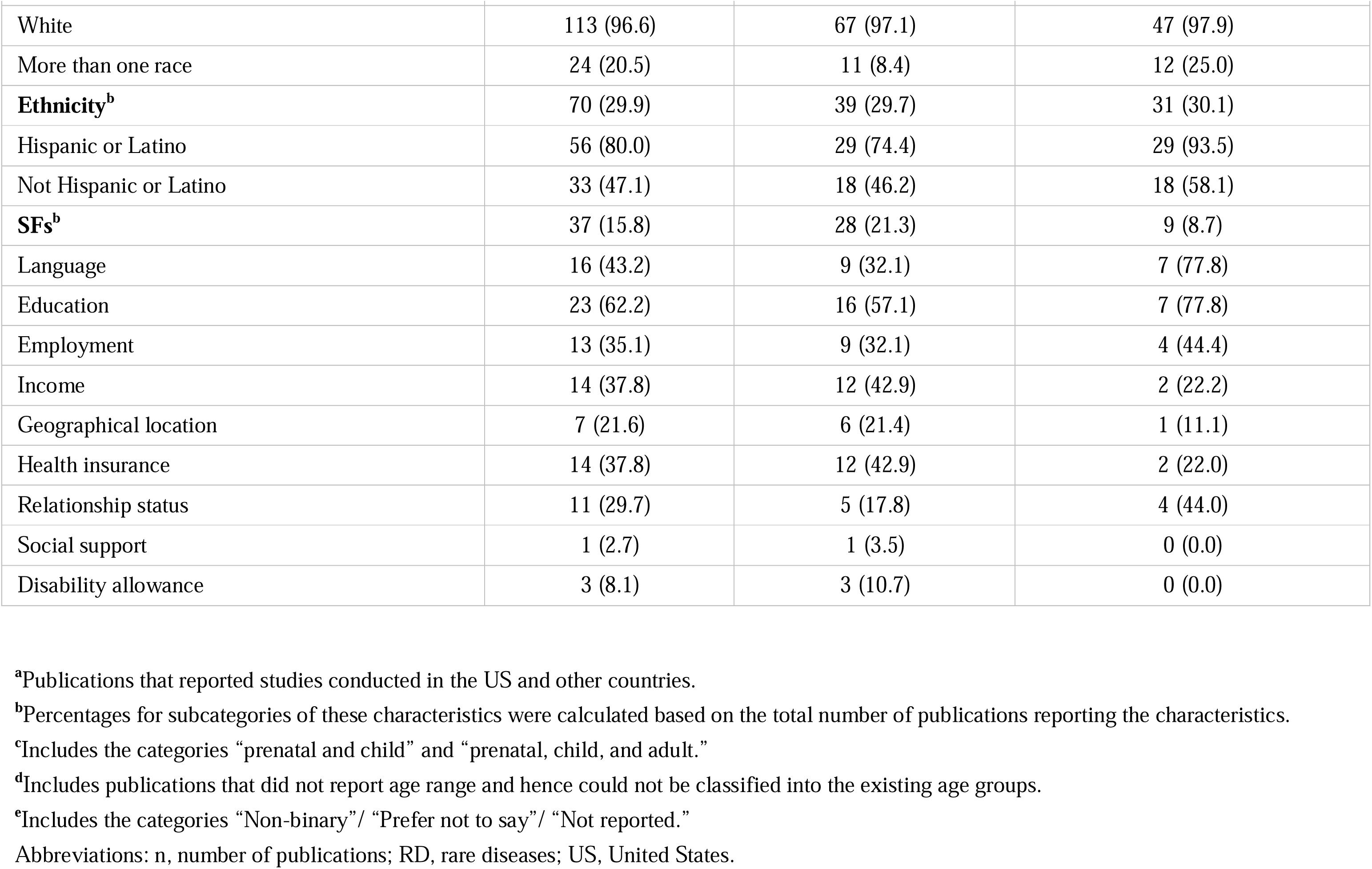
Characteristics of publications reporting RD research with US-based participants (2010–2023)

#### Demographic and SFs reporting in RD publications

The most reported demographic variable was age (94%), and the least reported was ethnicity (29.9%, Table 1). The most reported age category was “child and adult” (28.2%), followed by “adult and older adult” (25.5%), “child only” (14.2%), and prenatal (1.4%); “older adults” (0.9%) was the least reported category. Gender was reported by 86.3% of all publications; 66.8% of them reported it in the binary format. Only 5% of them reported a third option for gender as “other,” “non-binary,” “missing,” “not reported,” or “prefer not to say.” Race was reported in 50% of publications, and ethnicity in 29.9%; neither followed the standardized NIH/OMB format. Some publications used the term "Caucasian" for White participants, “non-White” for all other race participants, categorized race as ethnicity, reported data for some race categories, or combined data for two race categories (for example, Asian and Native Hawaiian or Other Pacific Islander were combined). In other publications, terms like “origin,” “ancestry,” or “country” were used to report race and ethnicity categories (Additional file 1: Table S4). Among the 117 publications reporting race, 96.6% included White participants, 65% Black or African American, 55.6% Asian, 21.4% American Indian or Alaska Native, 17.9% Native Hawaiian or Other Pacific Islander, and 20.5% more than one race. Hispanic and Latino participants were reported in 80% of the publications, which included ethnicity data (Table 1).

Only 37 of 234 (15.8%) publications reported one or more SFs; 75.7% of these were US-only publications. Education was the most reported SF (62.2%), and the least reported was social support (2.7%, Table 1). No standard reporting formats were applied; for example, education was reported in terms of the number of years or a categorical manner (“Bachelor,” “college,” “some college,” “high school”). Participants affected by RDs had varied educational backgrounds, insurance coverages, income levels, employment status, and relationship status, indicating potential inclusion and accessibility issues (Additional file 1: Table S5). English was the most reported language and was an inclusion/exclusion criterion in most publications reporting language data.

### ClinicalTrials.gov data analysis

From 44,064 US-based trials with study results, we identified 8,475 RD trials (1,492,423 participants; Figure S1B); 69.9% were US-only, and 30.1% were multinational trials (Table 2). 74% of the trials had a 100% keyword–condition match, and <5% of the trials were non-specific matches (Additional file 1: Table S6). Table 2 and Additional file 1: Table S7 summarize the trial characteristics, demographics reporting, and participant distributions.

**Table 2:**
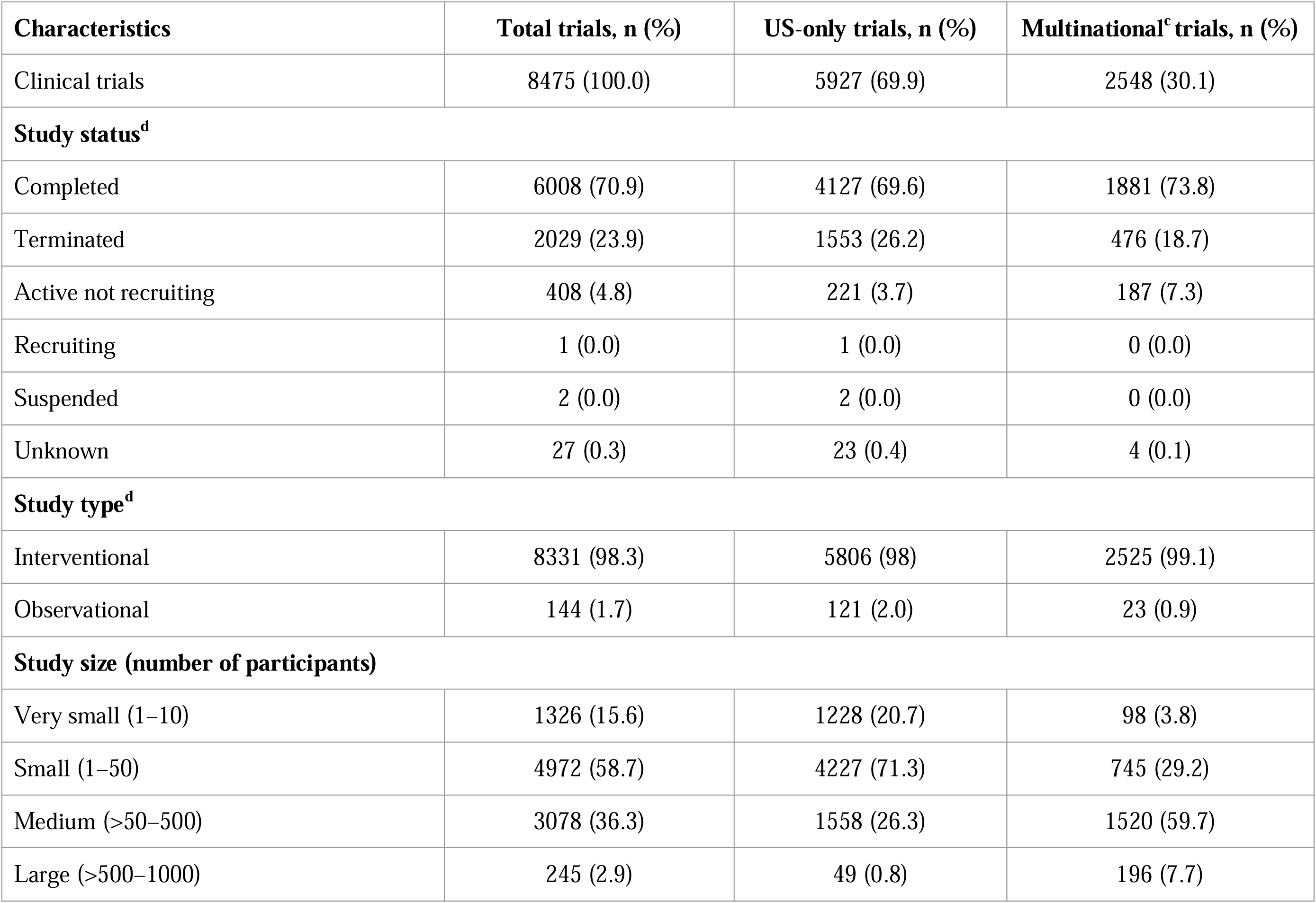
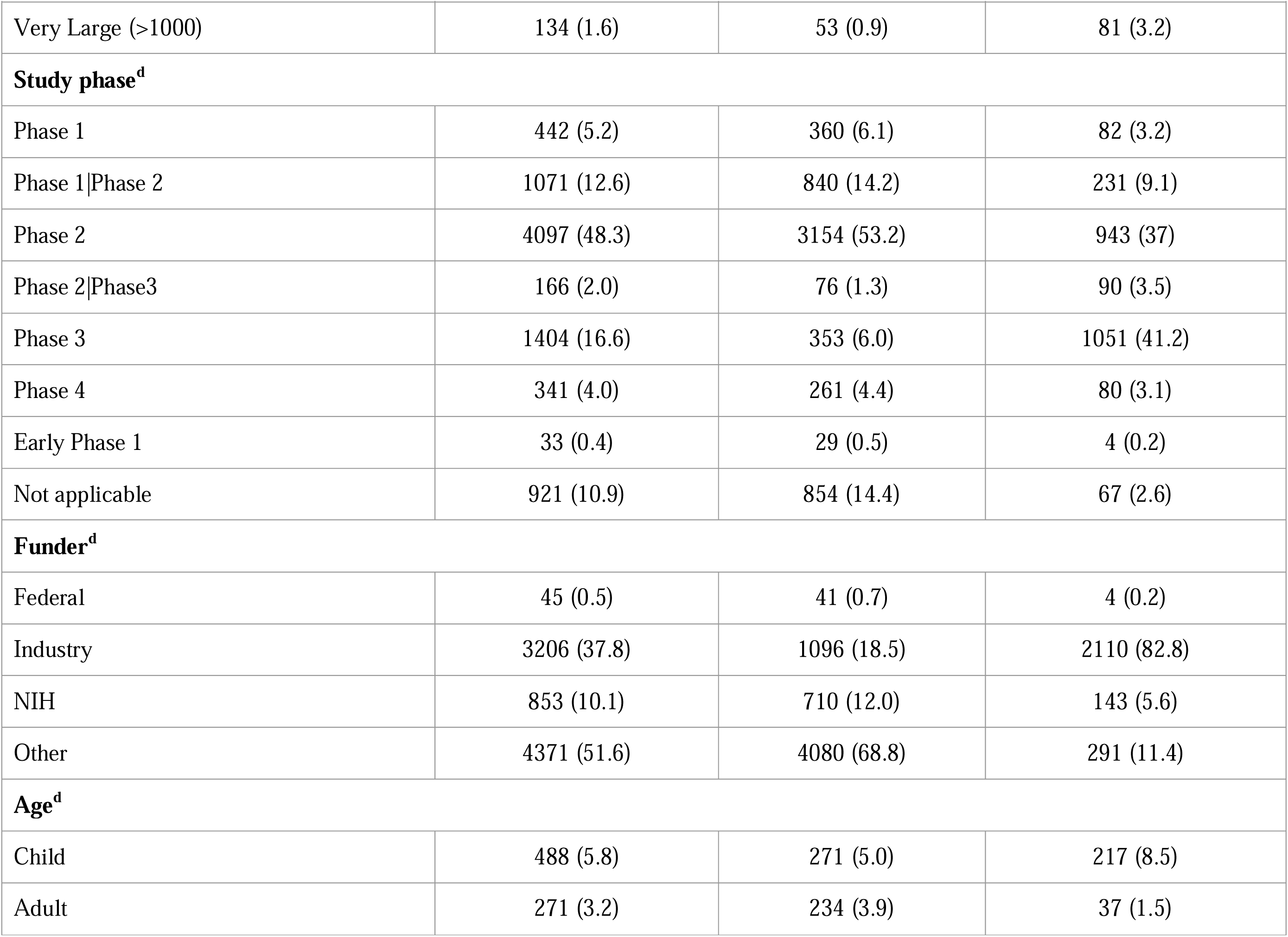
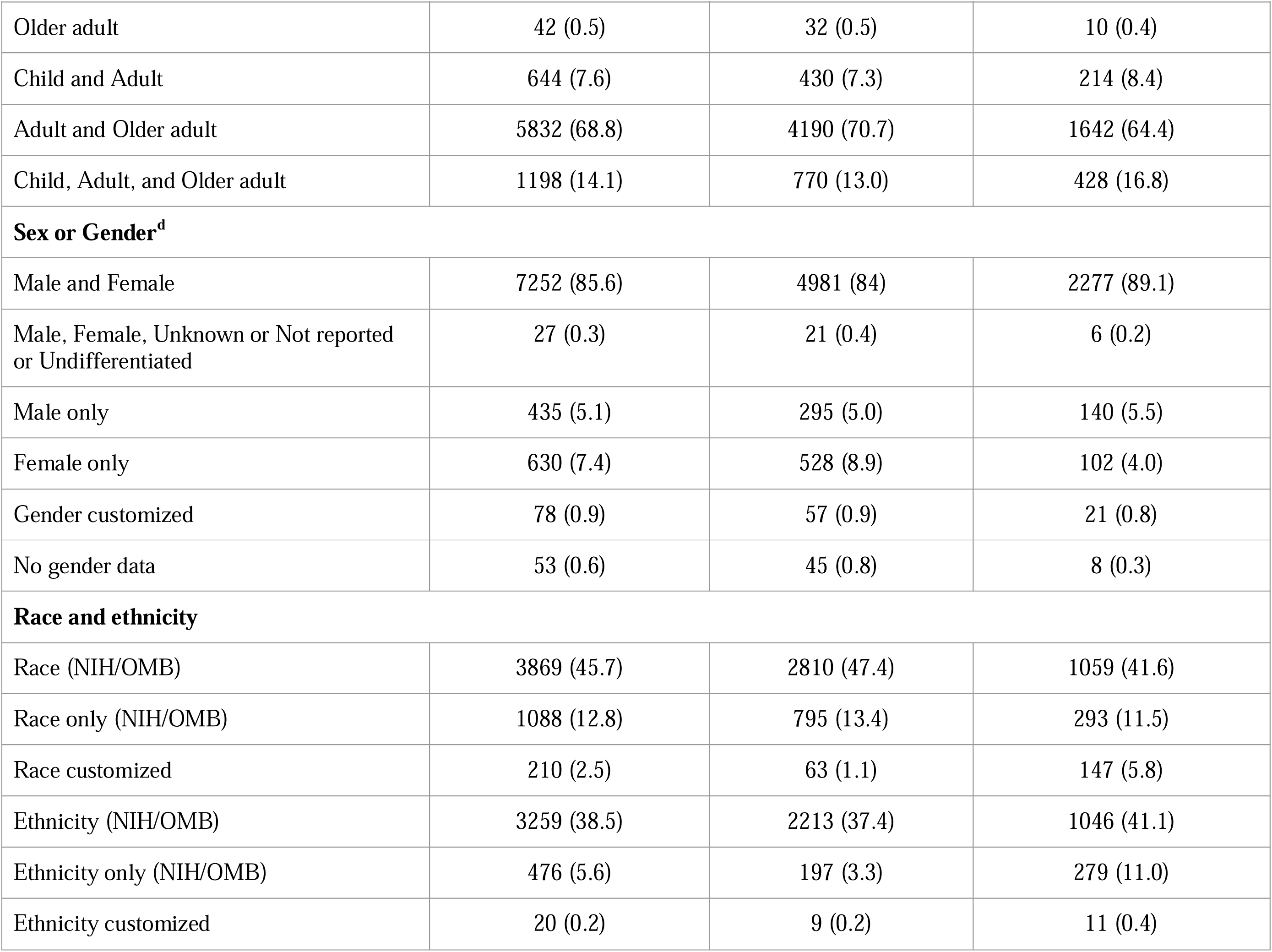
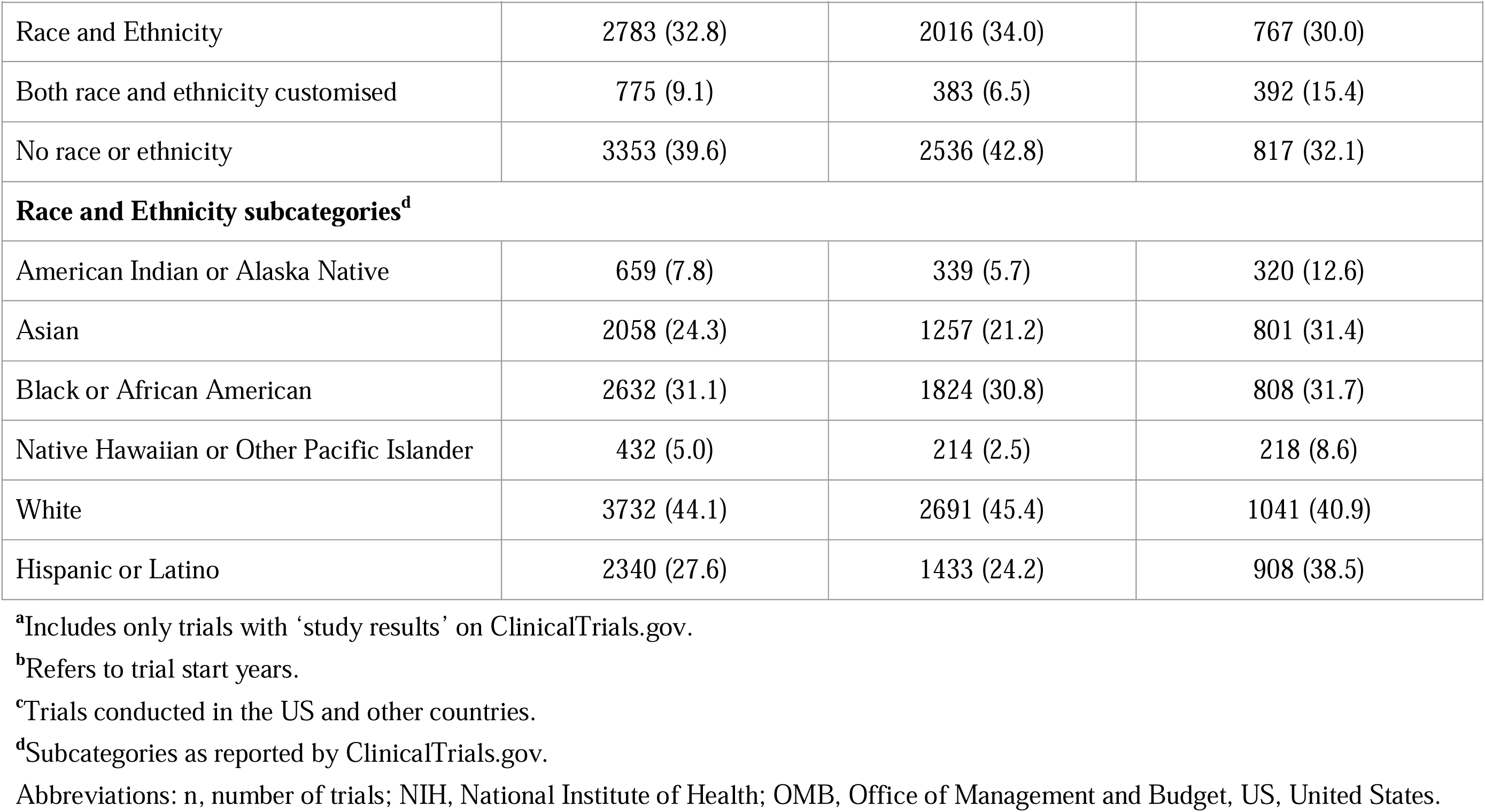
Characteristics of RDs clinical trials^a^ from ClinicalTrials.gov with US-based participants (1986–2022)^b^.

| Characteristics | Total trials, n (%) | US-only trials, n (%) | Multinational<sup>c</sup> trials, n (%) |
| --- | --- | --- | --- |
| Clinical trials | 8475 (100.0) | 5927 (69.9) | 2548 (30.1) |
| **Study status<sup>d</sup>** |  |  |  |
| Completed | 6008 (70.9) | 4127 (69.6) | 1881 (73.8) |
| Terminated | 2029 (23.9) | 1553 (26.2) | 476 (18.7) |
| Active not recruiting | 408 (4.8) | 221 (3.7) | 187 (7.3) |
| Recruiting | 1 (0.0) | 1 (0.0) | 0 (0.0) |
| Suspended | 2 (0.0) | 2 (0.0) | 0 (0.0) |
| Unknown | 27 (0.3) | 23 (0.4) | 4 (0.1) |
| **Study type<sup>d</sup>** |  |  |  |
| Interventional | 8331 (98.3) | 5806 (98) | 2525 (99.1) |
| Observational | 144 (1.7) | 121 (2.0) | 23 (0.9) |
| **Study size (number of participants)** |  |  |  |
| Very small (1–10) | 1326 (15.6) | 1228 (20.7) | 98 (3.8) |
| Small (1–50) | 4972 (58.7) | 4227 (71.3) | 745 (29.2) |
| Medium (>50–500) | 3078 (36.3) | 1558 (26.3) | 1520 (59.7) |
| Large (>500–1000) | 245 (2.9) | 49 (0.8) | 196 (7.7) |

| **Characteristics** | **Total trials, n (%)** | **US-only trials, n (%)** | **Multinational<sup>c</sup> trials, n (%)** |
| --- | --- | --- | --- |
| Very Large (>1000) | 134 (1.6) | 53 (0.9) | 81 (3.2) |
| **Study phase<sup>d</sup>** |  |  |  |
| Phase 1 | 442 (5.2) | 360 (6.1) | 82 (3.2) |
| Phase 1\|Phase 2 | 1071 (12.6) | 840 (14.2) | 231 (9.1) |
| Phase 2 | 4097 (48.3) | 3154 (53.2) | 943 (37) |
| Phase 2\|Phase3 | 166 (2.0) | 76 (1.3) | 90 (3.5) |
| Phase 3 | 1404 (16.6) | 353 (6.0) | 1051 (41.2) |
| Phase 4 | 341 (4.0) | 261 (4.4) | 80 (3.1) |
| Early Phase 1 | 33 (0.4) | 29 (0.5) | 4 (0.2) |
| Not applicable | 921 (10.9) | 854 (14.4) | 67 (2.6) |
| **Funder<sup>d</sup>** |  |  |  |
| Federal | 45 (0.5) | 41 (0.7) | 4 (0.2) |
| Industry | 3206 (37.8) | 1096 (18.5) | 2110 (82.8) |
| NIH | 853 (10.1) | 710 (12.0) | 143 (5.6) |
| Other | 4371 (51.6) | 4080 (68.8) | 291 (11.4) |
| **Age<sup>d</sup>** |  |  |  |
| Child | 488 (5.8) | 271 (5.0) | 217 (8.5) |
| Adult | 271 (3.2) | 234 (3.9) | 37 (1.5) |
| Older adult | 42 (0.5) | 32 (0.5) | 10 (0.4) |
| Child and Adult | 644 (7.6) | 430 (7.3) | 214 (8.4) |
| Adult and Older adult | 5832 (68.8) | 4190 (70.7) | 1642 (64.4) |
| Child, Adult, and Older adult | 1198 (14.1) | 770 (13.0) | 428 (16.8) |
| **Sex or Gender<sup>d</sup>** |  |  |  |
| Male and Female | 7252 (85.6) | 4981 (84) | 2277 (89.1) |
| Male, Female, Unknown or Not reported or Undifferentiated | 27 (0.3) | 21 (0.4) | 6 (0.2) |
| Male only | 435 (5.1) | 295 (5.0) | 140 (5.5) |
| Female only | 630 (7.4) | 528 (8.9) | 102 (4.0) |
| Gender customized | 78 (0.9) | 57 (0.9) | 21 (0.8) |
| No gender data | 53 (0.6) | 45 (0.8) | 8 (0.3) |
| **Race and ethnicity** |  |  |  |
| Race (NIH/OMB) | 3869 (45.7) | 2810 (47.4) | 1059 (41.6) |
| Race only (NIH/OMB) | 1088 (12.8) | 795 (13.4) | 293 (11.5) |
| Race customized | 210 (2.5) | 63 (1.1) | 147 (5.8) |
| Ethnicity (NIH/OMB) | 3259 (38.5) | 2213 (37.4) | 1046 (41.1) |
| Ethnicity only (NIH/OMB) | 476 (5.6) | 197 (3.3) | 279 (11.0) |
| Ethnicity customized | 20 (0.2) | 9 (0.2) | 11 (0.4) |
| Race and Ethnicity | 2783 (32.8) | 2016 (34.0) | 767 (30.0) |
| Both race and ethnicity customised | 775 (9.1) | 383 (6.5) | 392 (15.4) |
| No race or ethnicity | 3353 (39.6) | 2536 (42.8) | 817 (32.1) |
| **Race and Ethnicity subcategories<sup>d</sup>** |  |  |  |
| American Indian or Alaska Native | 659 (7.8) | 339 (5.7) | 320 (12.6) |
| Asian | 2058 (24.3) | 1257 (21.2) | 801 (31.4) |
| Black or African American | 2632 (31.1) | 1824 (30.8) | 808 (31.7) |
| Native Hawaiian or Other Pacific Islander | 432 (5.0) | 214 (2.5) | 218 (8.6) |
| White | 3732 (44.1) | 2691 (45.4) | 1041 (40.9) |
| Hispanic or Latino | 2340 (27.6) | 1433 (24.2) | 908 (38.5) |

<sup>a</sup>Includes only trials with ‘study results’ on ClinicalTrials.gov.
<sup>b</sup>Refers to trial start years.
<sup>c</sup>Trials conducted in the US and other countries.
<sup>d</sup>Subcategories as reported by ClinicalTrials.gov.
Abbreviations: n, number of trials; NIH, National Institute of Health; OMB, Office of Management and Budget, US, United States.

#### Reporting of demographics and SFs in RD trials

Most US-based RD trials included “adult and older adult participants” (68.8%) and both male and female participants (85.6%). Trials with only children and older adults accounted for 5.8% and 0.5%, respectively. Only 0.3% of the trials included non-binary sex or genders and reported them as unknown. Race was reported in 45.7% of the trials, ethnicity in 38.5% in the NIH/OMB format, while 9.1% used a customized format for reporting race and ethnicity. SFs were reported in 16 of 8475 (0.2%) trials; relationship status was most reported (68.8%), followed by education (50%), insurance status (37.5%), and the least reported was religion (6.3%, Additional file 1: Table S8 and S9).

#### Race and ethnicity in US-only RD trials

##### Participant-level analysis

US-only trials (1991–2022) reported race in 2,799 trials and ethnicity in 2,204 trials in the NIH/OMB format. Figure 1A summarizes racial and ethnic representation levels by presenting aggregate or overall proportions and mean proportions (the average of proportions from all trials) of participants. These aggregate and mean proportions significantly differed from the US census averages (race: χ²=5477.9 and 5500.9; ethnicity: χ²=15491077.3 and 13595347.8, p < 0.001).

**Figure 1:**
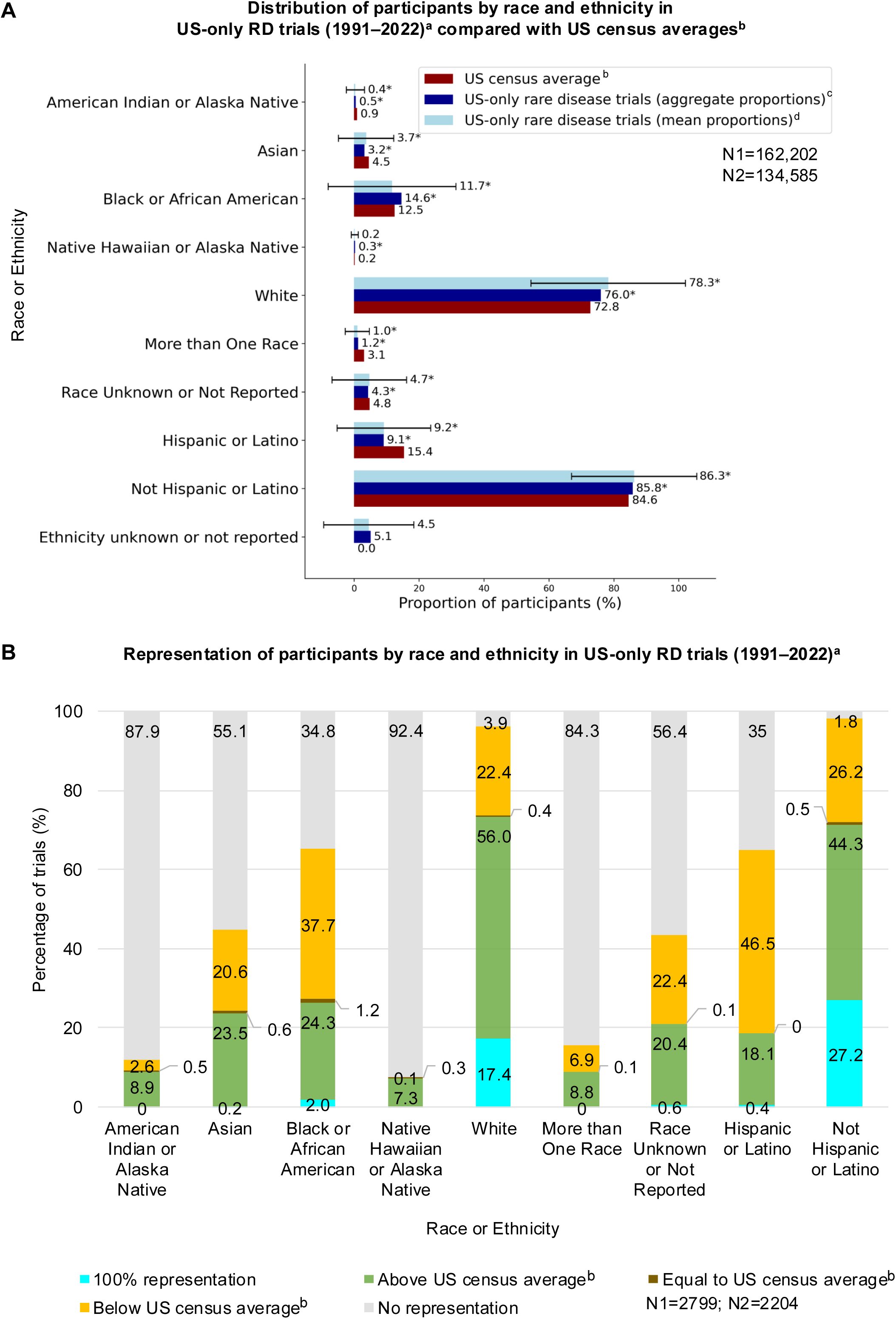
Distribution of participants by race and ethnicity: US-only RD trials (1991–2022)^a^ from ClinicalTrials.gov. **Figure 1A Caption:** Aggregate and Mean proportions of participants by race and ethnicity were compared with US census averages using a Chi^2^ test. Error bars for mean proportions indicate standard deviation. The significance of the “Ethnicity unknown or not reported” subcategory could not be evaluated as the US census lacks a corresponding category. **Figure 1A Legend:** ^a^Refers to the trial start years. **^b^**Weighted average percentages of US census data (1990–2022) calculated using weights based on the distribution of trial participants over time. **^c^**Aggregate proportions: calculated by dividing the number of participants of a racial or ethnic category by the total number of participants for whom race or ethnicity data are available. **^d^**Mean proportions: calculated by averaging the racial or ethnic proportions in each trial. *Indicates significance at Bonferroni-corrected level (race: 0071; ethnicity: 0167); *p-values* for significance in Table S10. Abbreviations: N1, total number of participants analyzed for race; N2, total number of participants analyzed for ethnicity; RD, rare disease; US, United States. **Figure 1B Caption:** The percentage of trials having representation for each race and ethnicity subcategory was evaluated by comparing aggregate proportions of participants with US census averages. “No representation” refers to trials with 0% aggregate proportions of participants for each race or ethnicity subcategory. The percentage of trials for representation of the“Ethnicity unknown or not Reported” subcategory could not be evaluated because the US census lacks a corresponding category. Data labels with category. **Figure 1B Legend:** ^a^Refers to trial start years. **^b^**Weighted average percentages of US census data (1990–2022) calculated using weights based on the distribution of trial participants over time. Abbreviations: N1, number of trials with study results reporting race data in NIH/OMB format; N2, number of trials with study results reporting ethnicity data in NIH/OMB format; RD, rare disease; US, United States.

Both aggregate ad mean proportions showed that the representation of American Indian or Alaska Native, Asian, and Hispanic or Latino participants was significantly below the US census averages; White representation was significantly above the US census average (Figure 1A, Additional file 1: Table S10). While aggregate proportions of Black or African American and Native Hawaiian or Pacific Islander participants were significantly above the US census averages, the mean proportion for Black or African American participants alone revealed the opposite trend. Across aggregate and mean proportions, 1–1.2% of the participants were categorized as more than one race, and 4.3–4.7% of the participants were “race unknown or not reported”; both were significantly below the US census averages. About 4.5–5.1% of the participants were “ethnicity unknown or not reported” with no corresponding category in the US census. (Figure 1A, Additional file 1: Table S10).

##### Trial-level analysis

We analyzed the trial-level diversity by assessing the percentage of trials having racial and ethnic representation above or below the US census average (Figure 1B). Most trials lacked representation for Native Hawaiian or Pacific Islander participants (92.4%), followed by American Indian or Alaska Native (87.9%), Asian (55.1%), Black or African American (34.8%), and Hispanic or Latino (35%) participants. In contrast, only 3.9% of the trials did not include White participants.

##### Study phase and funder type analysis

Across study phases and funder types, aggregate and mean proportions for race and ethnicity significantly differed from the US census averages (Figure 2, Additional file 1: Table S11). Only Black or African American, and White representation significantly exceeded the US census averages in at least one study phase or funder type (Figure 2, Additional file 2: Table S12). White representation significantly exceeded the US census average levels across funder types and study Phases 1–4. Conversely, Black or African American representation significantly exceeded US census average levels only in NIH-funded and Phase 1, 3, and 4 trials. Hispanic or Latino representation was lower than the US census average across study phases and funder type (Figure 2). The highest representation of Hispanic or Latino participants was in industry-funded trials (10.6–11.4%) and Phase 2|Phase 3 trials (12.1–15.5%).

**Figure 2:**
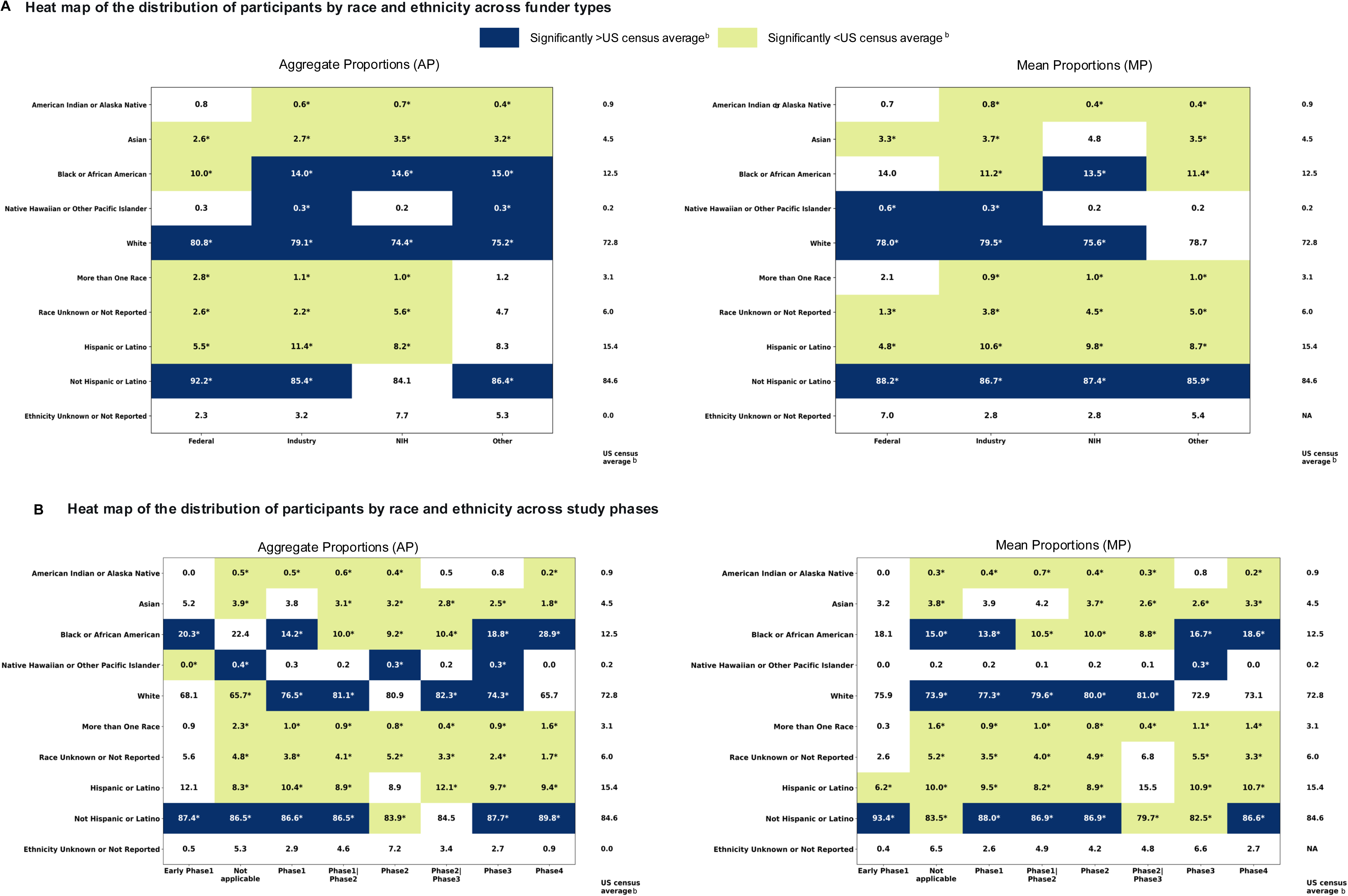
Participants distribution by funder type and study phase in US-only RD trials (1991–2022)^a^. **Figure 2 Caption:** Aggregate and mean proportions by race and ethnicity were calculated and compared with the US census averages^b^ using Chi^2^ tests. Post hoc analyses were conducted to determine the specific racial or ethnic groups contributing to the observed differences. Bonferroni correction was used to adjust the significance threshold. Proportions that significantly exceeded the US census averages^b^ are shaded in blue. **Figure 2 Legend:** ^a^Refers to trial start years. **^b^**Weighted average percentages of US census data (1990–2022) calculated using weights based on participant distribution in RD trials (1991–2022). *Indicates significance at Bonferroni-corrected significance level (AP: 0071; MP: 0063; MP funder type: 0125, *p-values* for significance in Table S11. Abbreviations: AP, aggregate proportion; MP, mean proportion; N, number of trials; NA, not available; RD, rare disease; US, United States.

##### Temporal trends

The number of RD clinical trials reporting race and ethnicity began increasing in 1999 (race: N_1_=8, ethnicity: N_2_=8) and peaked in 2015 for race (N_1_=282) and in 2016 for ethnicity (N_2_=230, Additional file 1: Figure S2A). We analyzed participant distribution over three decades (1991–2020) to better understand representation trends of the different racial and ethnic groups (Figure 3A–F, Additional file 1: Figure S3A–D). American Indian or Alaska Native, Asian, and Hispanic or Latino participation consistently remained below the US census. In contrast, Black or African American, and Native Hawaiian participation consistently remained higher than the US census for those three decades. White participation exceeded the US census for the decades between 2001–2020 (Figure 3).

**Figure 3:**
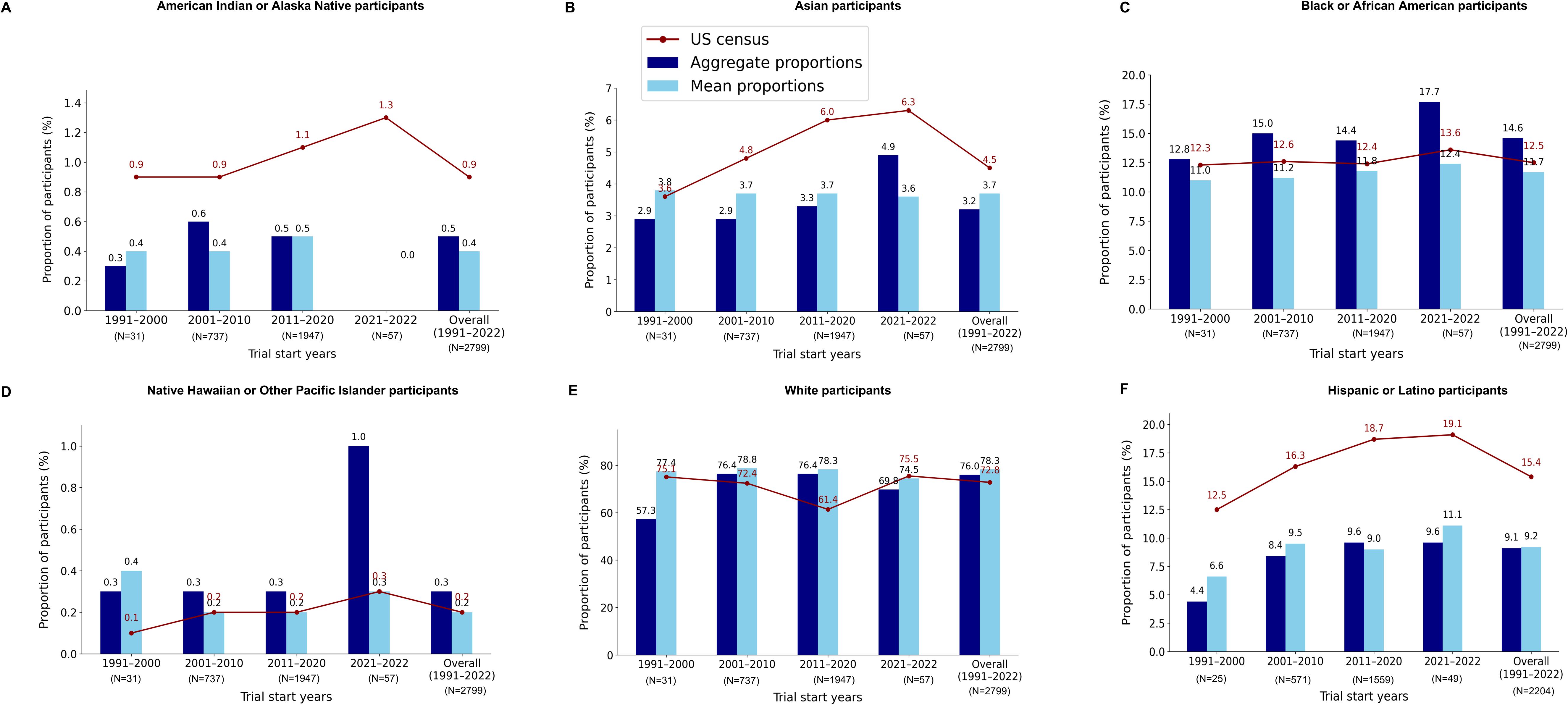
Temporal participation trends by race and ethnicity: US-only RD trials (1991–2022)^a^ from ClinicalTrials.gov. **Figure 3 Caption:** Aggregate and mean proportions by race and ethnicity were calculated for three decades (1991–2000, 2001–2010, and 2011–2020), the years 2021–2022, and the entire 30-year period (1991–2022). US census data for the years 2000, 2010, 2020, 2022, and US census averages (1990–2020) are plotted as a red line graph for comparison. US census averages (1990–2020) refer to weighted average percentages of US census data calculated using weights based on participant distribution in RD trials (1991–2022). **Figure 3 Legend:** ^a^Refers to trial start years Abbreviations: N, number of trials; RD, rare disease; US, United States.

#### Race and ethnicity in multinational trials

Multinational trials (1997–2022) reported race in 1051 trials and ethnicity in 1040 trials. Additional file 2: Table S13 presents the overall racial and ethnic distribution across funder types and study phases.

## Discussion

Our results reveal key gaps in demographics and SF reporting practices in RD research, emphasizing the urgent need to include MUPs. Consistent with previous studies, age and sex or gender were well reported in our analysis (45,46). There was no reporting of sexual orientation and gender identity (SOGI) data. We note that the FDA has not included SOGI in the DAP 2024 (4), whereas a Health Resources and Services Administration mandate existed from 2016 (47).

Race and ethnicity were underreported despite the Final Rule for Clinical Trials 2017(48). A previous study also reported that only 43% of US-based trials had race/ethnicity data (23). Only 14.5% of the publications included child participants, compared to 5.8% of the trials (Table 1). The presence of genomics/genetics studies (12.4%) in our literature analysis, which often includes child participants, may explain this discrepancy. An earlier report showed that 20% of global RD trials had child participants (26), contrasting with our analysis of US-based trials (5.8%). Older adult participants were underrepresented in publications (0.9%) and clinical trials (0.5%). Although a direct comparison with the FDA’s DTS (2015–2019) RD clinical trial data (>65 years of age=24%) is not possible (15), the underrepresentation of older adults is evident, considering they comprise 17% of the US population (49). However, we note that older adult participants were included in other age categories: “adult and older adult” or “child, adult and older adult.”

Most publications did not report race and ethnicity in the NIH/OMB format, making it research. Our analyses of aggregate and mean proportions of race and ethnicity highlight the persistent underrepresentation of Asian, Native Hawaiian or Other Pacific Islander, American Indian or Alaska Native, and Hispanic or Latino participants (Figure 2), consistent with other studies on US-based trials (22,23). Despite significantly higher aggregate proportions than the US census, the lower mean proportions (Figure 1A) and the percentage of trials lacking Black or African American (34.8%) and Native Hawaiian or Other Pacific Islander (92.4%) participants (Figure 1B) indicate a significant gap in participant recruitment.

Hispanic or Latino participation was numerically higher in our study (9.1%) than in the FDA DTS report (6%). Conversely, Asian participation was numerically lower in our study (3.2%) than in the FDA DTS (9%), likely because we focused on US-only trials, whereas FDA DTS included global RD trials.^15^ In multinational trials, both Asian (10.4%) and Hispanic or Latino (12.1%) participation were numerically higher in our study (Additional file 2: Table S13) compared with the FDA DTS. Both the FDA DTS and our multinational studies included Asians at lower rates compared with their global representation of 59.1% (50).

Although race and ethnicity reporting improved over time (Additional file 1: Figure S2, Figure S3B and S3D), the representation of American Indian, Asian, and Hispanic or Latino participants remained below the US census levels (Figure 3A, 3B, and 3F). The trend of overrepresentation of Black or African American, Native Hawaiian or Other Pacific Islanders could indicate uneven recruitment efforts. Although the percentage of people reporting multiple races increased to 10.2% in 2010, multiracial participation remained lower in RD trials in the period 2011–2020 and thereafter (Additional file 1: Figure S3A). The decline in trials reporting race and ethnicity data from 2018 onwards, consistent with a previous study (23), is likely due to our focus on trials with study results. However, whether the COVID-19 pandemic influenced the number of completed trials with study results is unclear.

The underrepresentation of racial and ethnic MUPs observed in this study could be due to barriers in accessing diagnostics and healthcare, limited awareness of trials, and resource constraints (51–53). Moreover, the historical mistrust in healthcare, stemming from events like the Tuskegee Syphilis Study (54), also contributes to lower participation of certain racial/ethnic groups (55–57). Implicit and explicit bias among healthcare providers results in certain racial and ethnic groups not being invited to participate in clinical research (58). Language barriers also hinder the participation of speakers with limited English proficiency (59,60). In our analysis, we found that English proficiency was used as an inclusion/exclusion criterion in studies reporting language data.

SFs were massively underreported, consistent with a previous study evaluating such reporting in high-impact factor medical journals (61). The discrepancy in SFs reporting between publications (15.8%) and clinical trials (0.2%, Table 1) is likely due to the focus on clinical and biomedical data rather than SFs in ClinicalTrials.gov. A recent report showed that SFs in US clinical research sites are the least collected data (62). The limited or lack of data on veterans, immigrants, religious minorities, rural residents, and those affected by poverty is likely because these are not federally mandated data elements. Additionally, privacy and discrimination concerns could also contribute to the barriers to collecting data on veterans and immigrants.

### Limitations

This study has limitations. The literature search strategy used “RDs” or “rare disorders” in all fields and Medical Subject Headers (MeSH) terms. However, not all RD publications are annotated with these MeSH terms (63), causing dataset gaps. Most publications did not report race/ethnicity in a standardized format, and 30–50% omitted these data. Only publicly available trials with study results in ClinicalTrials.gov were analyzed, and 1005 trials with customized race/ethnicity were excluded. Racial and ethnic representation in trials were compared with the US census, which is unsuitable for individual RDs with differing prevalence across races and ethnicities. Ideally, representation should also be evaluated based on the epidemiology of the disease (24), consistent with FDA guidance (4). However, as our analysis included all RD trials, the US census was a practical choice for comparisons.

## Conclusions

This study is the first comprehensive report quantifying DEIA variables in US RD clinical research. We identified reporting gaps for race, ethnicity, and SFs in publications and clinical trials. There is a lack of data on several MUPs, coupled with the underrepresentation of racial and ethnic minority groups. Our observations support the urgent need for data-driven recommendations to improve measurement, reporting, and a national framework to enhance DEIA in RD research. Our study could serve as a template for future global studies evaluating DEIA variables in not just RD research but all clinical research.

## Data Availability

This study used publicly available data. The rare disease keywords, publications, and clinical trial data included in the analysis are available from the corresponding author upon reasonable request.

## Abbreviations

US: United States
FDA: Food and Drug Administration
DAP: Diversity Action Plan
RD: Rare Disease
DEIA: Diversity, Equity, Inclusion, Accessibility
DTS: Drug Trial Snapshot Summary
CF: Cystic Fibrosis
MUPs: Medically underserved populations
SFs: Socioeconomic Factors
MUAs: Medically underserved areas
USA: United States of America
GARD: Genetic and Rare Disease Information Centre
LGBTQ+: lesbian, gay, bisexual, transgender, queer or questioning
NIH: National Institute of Health
OMB: Office of Management and Budget
SOGI: Sexual Orientation and Gender Identity
MeSH: Medical Subject Heading

## Declarations

### Ethics approval and consent to participate

Not applicable

### Consent for publication

Not applicable

### Author contributions

HKR, LM, and SMS contributed to the conceptualization of the study. LM and SMS acquired data, developed methods, conducted investigation, validated the data, drafted the manuscript, and prepared figures; SMS created new software for data extraction. LM, DD, and SMS analyzed the data, and DD performed statistical analysis. LM, SMS, DD NV, RVK, HKR, JNW, and LGB interpreted the data, reviewed it for important intellectual content, and revised and edited the manuscript. HKR, NV, and RVK obtained funding for the study. LM managed the project administration with JNW and LGB, providing administrative, technical, or material support. HKR supervised the study. All authors read and approved the manuscript.

## Acknowledgments

IndoUSrare acknowledges the RDDC for providing funding. The authors thank the RDDC Clinical Trials Working Group members for their invaluable contributions during manuscript development. We acknowledge Ms Bethlehem Addisu Demissie, Ms Bethlehem Sisay Tefera, Ms Cindy Diana Umanzoar Figueroa, Dr Dora Akinyi Mugambi, and Ms. Harriet Tunu Baraka for partial literature screening; Dr Dora Akinyi Mugambi, Ms Leanne Marie Woehlke, Ms. Kathleen Machuzak, Ms Veronica Mullins, and Dr Paul I Howard for analysis, interpretation, and content review; Dr Debra S Regier for editorial support; and Ms. Jocelyn Cooper and Ms. Lolita Smith-Moore for administrative support. We also acknowledge support received from Children’s National Hospital. We also thank Dr Pragya Chaube for her initial involvement in the literature search strategy and IndoUSrare volunteers for their support in partial literature screening and data extraction: Dr Dhanya K, Ms Ramya Muralikrishna, Mr Rohan Honganoor, and Mr Vikram Nadathur. We also thank Dr Kirkland Wilson for his input on rare disease filtering and statistical tests.

## Authors’ information

Indo US Organization for Rare Diseases, Herndon, VA 20171, USA

Lavanyaa Manjunatha, PhD; Saundarya MS, MSc; Deepika Dokuru, PhD; Nisha Venugopal, PhD; Reena V Kartha, PhD; Harsha K Rajasimha MS, PhD

Center for Orphan Drug Research, Department of Experimental and Clinical Pharmacology, University of Minnesota Twin Cities, Minneapolis, MN, USA

Reena V Kartha, PhD

Jeeva Clinical Trials Inc., Manassas, VA 20109, USA; School of Systems Biology, George Mason University, Fairfax, VA 22030, USA

Harsha K Rajasimha MS, PhD

Rare Disease Diversity Coalition, Black Women’s Health Imperative, Atlanta, GA 3013, USA

Jenifer Ngo Waldrop, MS; Linda Goler Blount, MPH

## Additional information

### Additional File 1_Manjunatha et al_Current Trends_MUP State in RD research_16022025

Additional File 1: Supplementary Methods 1 and 2, Figures S1–S3, and Tables S1–S11. Supplementary Methods 1. Literature assessment screening and data extraction; Supplementary Methods 2. Rare disease clinical trials filtering and validation; Supplementary References; Figure S1: Screening and filtering of US-based RD publications and clinical trials from ClinicalTrials.gov; Figure S2: Temporal trends in US-only RD trials reporting race and ethnicity data over time; Figure S3: Temporal participation trends: More than one race, Not Hispanic or Latino, Race and Ethnicity unknown or not reported in US-only RD trials (1991–2022); Table S1: PubMed and Cochrane Library search queries; Table S2: Keywords used for title screening of publications; Table S3: Keywords for identifying RD clinical trials reporting socioeconomic factors data; Table S4: A snapshot of US-based RD publications reporting race and ethnicity in a non-standard format; Table S5: Socioeconomic factors reporting in US-based RD publications; Table S7: Participant distribution statistics from US-based RD clinical trials (1986–2022); Table S8: Socioeconomic factors reporting in US-based RD trials (1991–2022); Table S9: Participant distribution statistics in US-based RD trials reporting socioeconomic factors (1991–2022); Table S10: p-values for statistical significance of aggregate and mean race and ethnicity proportions, in US-only RD trials (1991–2022), compared with the US census averages; Table S11: p-values for statistical significance of aggregate and mean proportions of race and ethnicity in US-only RD trials (1991–2022), compared with US census averages: by study phase and funder type

### Additional File 2_Table S12 and S13_Manjunatha et al_Current State_MUP trends in RD research_1602025.xlsx

Additional File 2: Table S12–S13. Table S12: p-values for significance of aggregate and MP of race and ethnicity in US-only RD trials (1991– 2022) compared with US census averages: by study phase and funder type; Table S13: Racial and ethnic distribution of participants in multinational RD trials (1997–2022)

