## Supplementary methods, figures, tables for "Current State and Demographic Trends of Medically Underserved Populations in Rare Disease Research in the United States"

### Supplementary Methods 1. Literature assessment screening and data extraction

#### Screening and Filtering

The literature search queries included terms (Table S1) from the definition of underserved communities (Box 1). Publication titles, grouped by database, were deduplicated by title and PubMed Central Identifiers (PMCIDs). Full-text publications were filtered by PMCID, PubMed Identifier, or Digital Object Identifier. Titles were programmatically and manually screened for specific keywords (Table S2). The resulting set was manually screened by title and abstract by evaluating whether they reported one or more rare disease (RD) conditions, had study location as the US, and reported participant demographics or socioeconomic factors.

Furthermore, publications older than 2010 were filtered out. During the full-text screening, the country location of participants was determined by examining the methods and results section or the supplementary section. In the absence of this data, we consulted the associated clinical trial if a National Clinical Trial (NCT) number was provided in the publication. Further, if NCT numbers were unavailable, we examined the author affiliations and assessed the location information. In the final full-text screening, studies were included in the analysis only if they satisfied all inclusion criteria. Case reports or clinical reports were excluded if they had demographics or SF data for only one participant.

#### Data extraction

The extracted data items included disease condition, location, age, sex or gender, race, ethnicity, language, education, employment, income, residence location, health insurance, relationship status, disability allowance, and social support (special needs assistance and food assistance). Age data were categorized as follows: prenatal (gestational weeks), child (0 months–17 years), adult (18–65 years), and older adult (≥65 years) based on the ranges reported in the Methods, Tables, Results, or Supplementary section of the publication. We recognize that sex and gender are distinct concepts; however, in this analysis, we refer to “sex or gender” as many publications did not specify whether the reported data were for sex or gender. For publications that reported data for only one sex or gender, it was assumed that the remaining data corresponded to the binary counterpart. Data labeled as Caucasian or Non-Hispanic White were categorized as White, while data labeled as Non-Hispanic Black were categorized as Black. If data for other races were explicitly mentioned, they were placed in the ‘other’ category. When data for two races were combined, they were categorized as ‘one or more races.’ When only one race category was reported, it was assumed that other categories were not reported, and the data were computed as 100% minus the reported race data. All publications reporting sex or gender data were also examined for keywords such as lesbian, gay, bisexual, transgender, queer, gender non-conforming, and non-binary (LGBTQ+). Publications reporting social factors (SF) were investigated for veterans, military spouses, religion, and immigrants, as the extracted SF data elements would not directly provide information about these marginalized urban populations (MUPs).

**Data synthesis**

Descriptive statistics were used to report the number of publications reporting each demographic and SF data and their subcategories. SF data were extracted as reported in the publications and synthesized based on the sub-categorizations for each type of SF along with the range of participant proportions to get a broad overview of representation. As SF data were variably reported, AI assistance (ChatGPT-4 Turbo, Open AI) was used to create standardized bins for the subcategories. We input the manually extracted subcategories and the corresponding participant proportions into ChatGPT3.5 and instructed it to develop standardized bins. These were manually verified for accuracy. Further, we asked ChatGPT to organize the manually extracted individual data points into the respective bins and extract the minimum and maximum participant proportions for each bin; we manually verified all AI outputs and created Table S5.

### Supplementary Methods 2. Rare disease clinical trials filtering and validation

#### Compilation of rare disease terms

We compiled rare disease terms from the Genetic and Rare Diseases Information Center (GARD), Orphanet, Rare-X, and ClinicalTrials.gov. ^1–4^ The GARD list included several comma-separated synonyms; these were extracted and added to the main list. After removing duplicates using Excel, we generated a final list of 30,303 unique keywords.

#### Initial keyword matching

We then performed a non-stringent matching of these keywords against the conditions field in the 44,064
US-based clinical trial data (study information downloaded in CSV) and Levenshtein distance scoring were used to obtain a similarity percentage, resulting in 39,095 unique NCT numbers. About 29,000 matches to non-rare disease conditions non-specifically matched to acronyms were eliminated.

#### Addressing non-specific acronym matches

About 10% of the filtered NCTs were non-specifically matched to acronyms, often due to word order and the presence of commas, hyphens, and other special characters. To address this issue, we performed normalized and tokenized matching using the non-stringent matching script, normalized matching using a regex matching pattern, and reordered matching of comma-separated conditions.

#### Final filtering process

We conducted another non-stringent matching with the remaining 34,000 NCTs for final filtering, ensuring all NCTs matched with non-acronyms had a Levenshtein similarity score above 50%. As Rare-X includes several keywords sourced from Online Mendelian Inheritance in Man known to have rare genetic variants, NCTs matched with keywords from Rare-X were included only when the prevalence was less than 200,000. Additionally, we mapped the matched keywords back to our rare disease list to identify those keywords that were only sourced from a single database, for example, Rare-X, GARD, or the rare disease list from ClinicalTrials.gov. We checked for prevalence data for such disease terms and included only those with a prevalence >200,000 or if no prevalence data were available.

#### Manual verification

We automatically included all NCTs with 80–100% similarity scores and performed manual verification of all NCTs with a similarity score below 80%. Of the 8,475 trials, 74.0% of the NCTs had a 100% keyword–condition match. Matches below 50% accounted for 7.6% of NCTs, while non-specific acronym matches accounted for 4.2% of NCTs.

#### Data compilation and verification

A master data set was created, including the trials from 1983–2023. Data extracted included study status, study phase, study type, sponsor type, brief title, start year, countries, region of enrolment, participant count, age, gender, race, and ethnicity. All data were extracted from the downloaded XML files, except age data, which were extracted from the study information (CSV files). Data completeness was verified by ensuring all fields contained valid entries. Participant counts were matched with the gender, race, and ethnicity counts. For those trials where these data did not match, the NCTs were revisited to understand the participant flow. Trials were classified as ‘complex’ when the participant counts could not be reconciled owing to overlap in participants in treatment arms, the study being a multipart one, or if the data were not reported for participants (for example, a trial reported the number of eyes receiving treatment).

#### Analysis

Trials with ‘complex’ data were excluded from any analysis of participant distributions. Trials where sex or gender were customized were excluded from sex or gender participant distribution calculations. Only trials reporting race/ethnicity in the NIH/OMB format were included in the race analysis. Aggregate and mean proportions of participants were calculated to assess racial and ethnic representation levels. Aggregate proportions were calculated by dividing the number of participants belonging to a particular race/ethnicity subcategory by the total number of participants for whom race/ethnicity was reported. As aggregate proportions are direct calculations, there were no measures of variability. Mean proportions were calculated by averaging the racial and ethnic proportions over all trials, and standard deviations were calculated. For comparing the representation with the US population, census data were downloaded for 1990, 2000, 2010, 2020, and 2022^5–7^. For an accurate comparison of trial representation and population representation, weighted averages were calculated based on the aggregate participant distribution in the clinical trials for the census bins.

### Figure S1: Screening and filtering of US-based RD publications and clinical trials from ClinicalTrials.gov

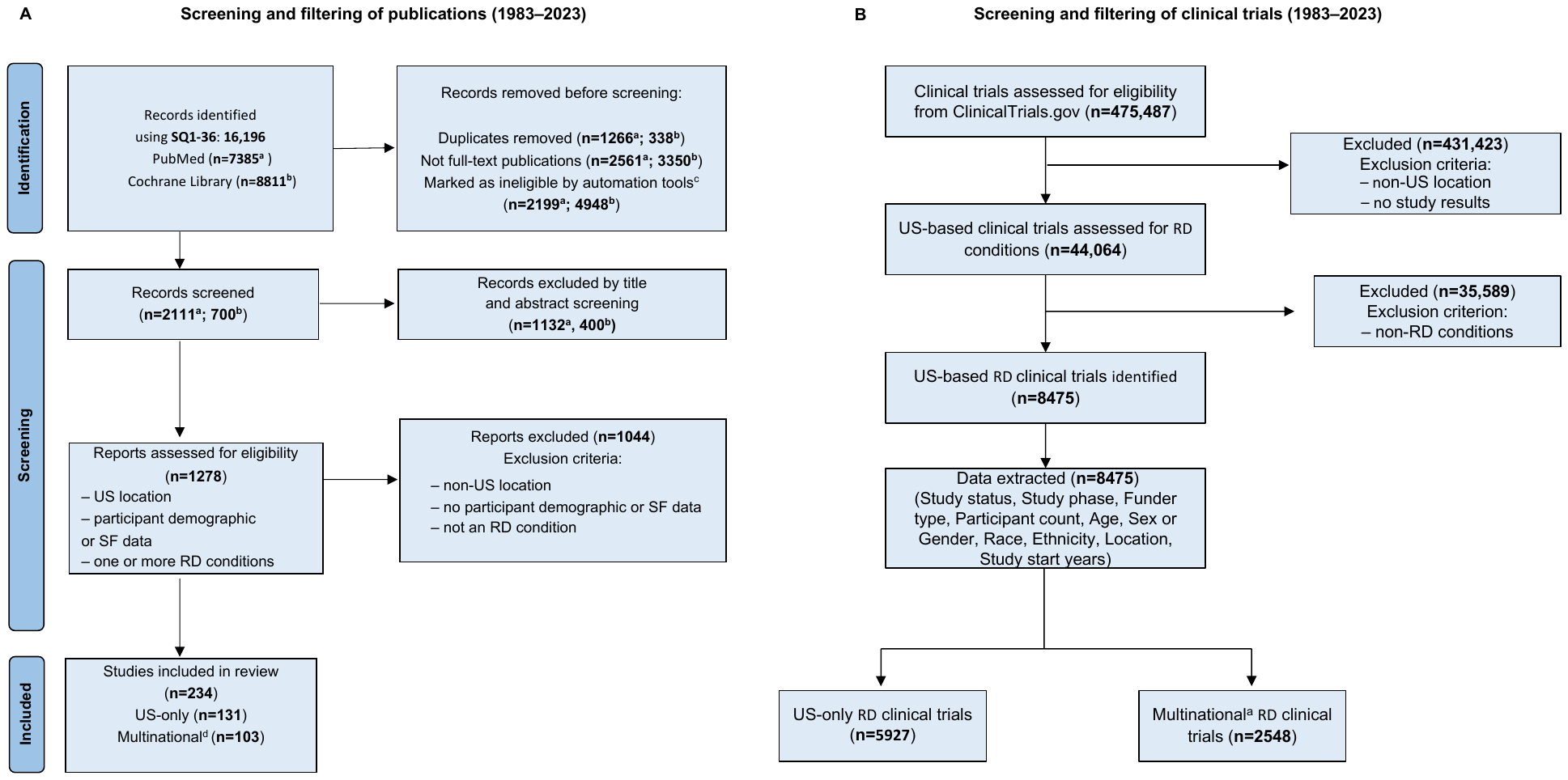

##### Figure S1a Caption and Legend: Search queries (SQ1–36, Table S1) were used to search PubMed and Cochrane Library. After deduplication and excluding publications without a full text, a combination of automation and manual screening of keywords (Table S2) was performed, followed by a title and abstract screening. Full-text screening assessed if publications met the inclusion criteria. The final set of publications included in the analysis were from 2010 to 2023.

##### ^a^publications from PubMed; ^b^publications from Cochrane Library; ^c^screened by the absence of one or more keywords as in Table S2; ^d^includes US and other countries; n, number of publications; SF, socioeconomic factors; SQ, search queries.

##### Figure S1b legend: ^a^Trials conducted in the US and other countries; n, number of clinical trials.

### Figure S2: Temporal trends in US-only RD trials reporting race and ethnicity data over time

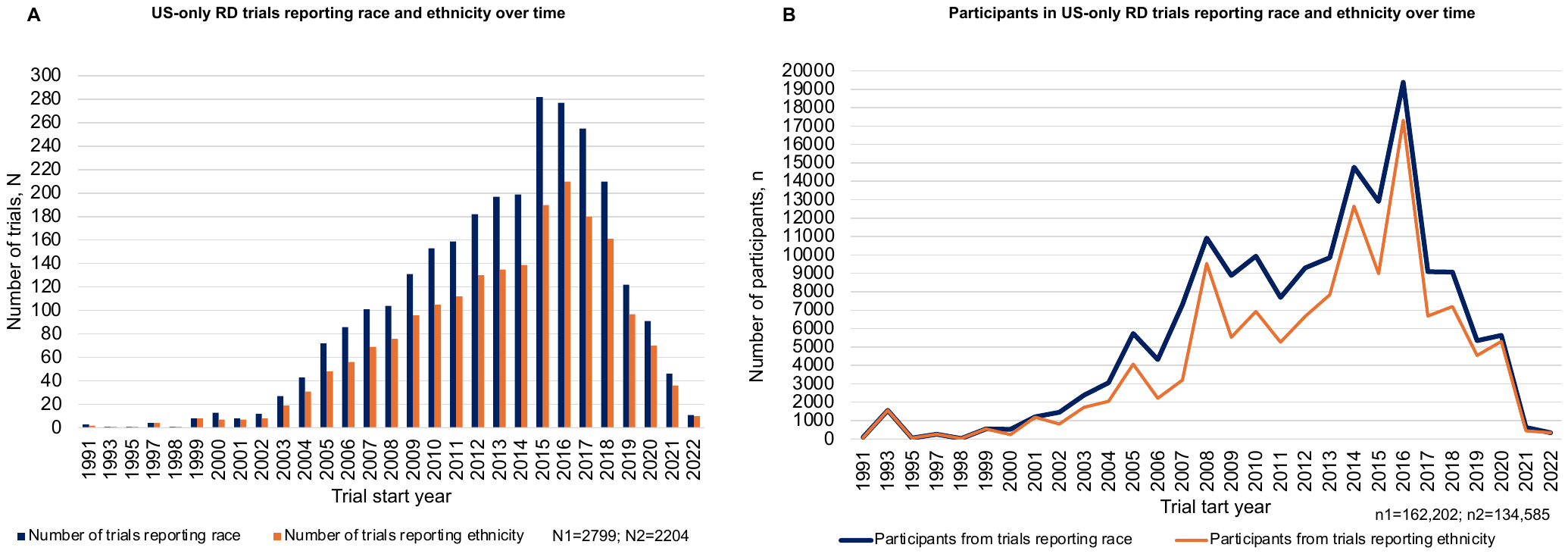

N1, number of trials reporting race; N2, number of trials reporting ethnicity; n1, number of participants for whom race was reported; n2, number of participants for whom ethnicity was reported; RD, rare disease; US, United States.

### Figure S3: Temporal participation trends: More than one race, Not Hispanic or Latino, Race and Ethnicity unknown or not reported in US-only RD trials (1991–2022)^a^

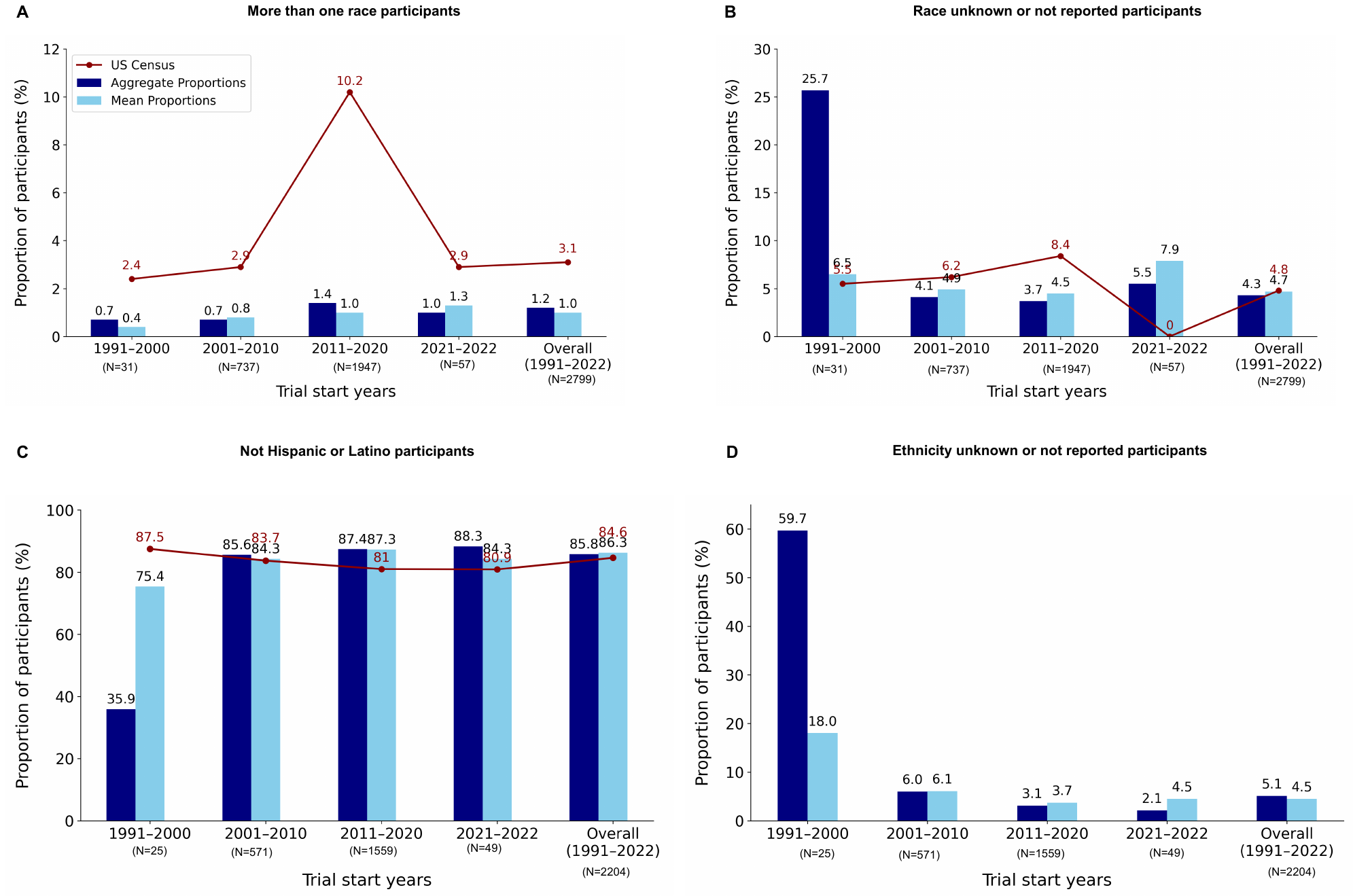

##### Aggregate and mean proportions by race and ethnicity were calculated for three decades (1991–2000, 2001–2010, and 2011–2020), the years 2021–2022, and the entire 30-year period (1991–2022). The US census data for the years 2000, 2010, 2020, and 2022 and the US census averages (1990–2020) are plotted in red as a line graph for comparison. Panel D does not have a US census data plotted as there is no corresponding category in the census.

##### ^a^Refers to trial start years; N, number of trials; RD, rare disease; US, United States.

### Table S1: PubMed and Cochrane Library search queries

| Search query (SQ) number | Search query string | Database | Number of results |
| --- | --- | --- | --- |
| SQ 1 | ("Rare Diseases"[MeSH] OR "Rare condition"[MeSH] OR "Rare disorder"[MeSH] OR "Orphan diseases"[MeSH] OR "Neglected disease"[MeSH]) AND (("1983/01/01"[PDAT]: "2023/06/30"[PDAT]) AND "humans"[MeSH Terms] AND English[lang] AND ("USA" OR "United States" OR "America")) | PubMed | 134 |
| SQ 2 | SQ2: ("Rare Diseases"[MeSH] OR "Rare condition"[MeSH] OR "Rare disorder"[MeSH] OR "Orphan diseases"[MeSH] OR "Neglected disease"[MeSH]) AND (("1983/01/01"[PDAT]: "2023/06/30"[PDAT]) AND "humans"[MeSH Terms] AND English[lang] AND ("USA" OR "United States" OR "America") AND ("diversity" OR "DEIA" OR "Inclusion" OR "underrepresented" OR "socioeconomic" OR "race" OR "gender" OR "LGBTQ" OR "queer" OR "disability" OR "ethnicity") | PubMed | 14 |
| SQ 3 | ("Rare Diseases"[MeSH] OR "Rare condition"[MeSH] OR "Rare disorder"[MeSH] OR "Orphan diseases"[MeSH] OR "Neglected disease"[MeSH]) AND (("1983/01/01"[PDAT]: "2023/06/30"[PDAT]) AND "humans"[MeSH Terms] AND English[lang] AND ("USA" OR "United States" OR "America") AND ("diversity" OR "DEIA" OR "Inclusivity") | PubMed | 2767 |
| SQ 4 | ("Rare Diseases"[MeSH] OR "Rare condition"[MeSH] OR "Rare disorder"[MeSH] OR "Orphan diseases"[MeSH] OR "Neglected disease"[MeSH]) AND (("1983/01/01"[PDAT]: "2023/06/30"[PDAT]) AND "humans"[MeSH Terms] AND English[lang] AND ("USA" OR "United States" OR "America") AND ("Clinical trials" OR "genomics" OR "whole-genome" OR "gene editing" OR "CRISPR" OR "transcriptomics" OR "human genome")) | PubMed | 554 |
| SQ 5 | ("RareDisease" OR "Rare Disease " OR "Rare Disorder ") AND ("Diversity" OR "Demographics" OR "Epidemiology") AND ("Randomised control trials" OR "Clinical trials") AND ("USA" or "United States of America" OR "America") | PubMed | 294 |
| SQ 6 | Rare disease OR rare disorder OR rare genetic disorder in All Text AND USA in All Text AND English Language | Cochrane | 8813 |
| SQ 7  Underserved | (("rare diseases"[All Fields] OR "rare diseases"[MeSH Terms] OR "rare condition"[All Fields] OR "rare disorder"[All Fields] OR "orphan diseases"[All Fields] OR "neglected diseases"[All Fields]) AND ("underserved"[All Fields] OR "underserviced"[All Fields] OR "underservicing"[All Fields])) AND ((ffrft[Filter]) AND (humans[Filter]) AND (1983/1/1:2024/4/12[PDAT]) AND (english[Filter])) | PubMed | 23 |
| SQ8  pregnancy | (("rare diseases"[All Fields] OR "rare diseases"[MeSH Terms] OR "rare condition"[All Fields] OR "rare disorder"[All Fields] OR "orphan diseases"[All Fields] OR "neglected diseases"[All Fields]) AND ("pregnancy"[MeSH Terms] OR "pregnancy"[All Fields] OR "pregnancies"[All Fields] OR "pregnancy s"[All Fields])) AND ((ffrft[Filter]) AND (humans[Filter]) AND (1983/1/1:2024/4/12[PDAT]) AND (english[Filter])) | PubMed | 922 |
| SQ9  caregivers | (("rare diseases"[All Fields] OR "rare diseases"[MeSH Terms] OR "rare condition"[All Fields] OR "rare disorder"[All Fields] OR "orphan diseases"[All Fields] OR "neglected diseases"[All Fields]) AND ("caregiver s"[All Fields] OR "caregivers"[MeSH Terms] OR "caregivers"[All Fields] OR "caregiver"[All Fields] OR "caregiving"[All Fields])) AND ((ffrft[Filter]) AND (humans[Filter]) AND (1983/1/1:2024/4/12[PDAT]) AND (english[Filter])) | PubMed | 245 |
| SQ10  parents | (("rare diseases"[All Fields] OR "rare diseases"[MeSH Terms] OR "rare condition"[All Fields] OR "rare disorder"[All Fields] OR "orphan diseases"[All Fields] OR "neglected diseases"[All Fields]) AND ("parent s"[All Fields] OR "parentally"[All Fields] OR "parentals"[All Fields] OR "parented"[All Fields] OR "parenting"[MeSH Terms] OR "parenting"[All Fields] OR "parents"[MeSH Terms] OR "parents"[All Fields] OR "parent"[All Fields] OR "parental"[All Fields])) AND ((ffrft[Filter]) AND (humans[Filter]) AND (1983/1/1:2024/4/12[PDAT]) AND (english[Filter])) | PubMed | 623 |
| SQ11  disability | (("rare diseases"[All Fields] OR "rare diseases"[MeSH Terms] OR "rare condition"[All Fields] OR "rare disorder"[All Fields] OR "orphan diseases"[All Fields] OR "neglected diseases"[All Fields]) AND ("disabilities"[All Fields] OR "disability"[All Fields] OR "disabled persons"[MeSH Terms] OR ("disabled"[All Fields] AND "persons"[All Fields]) OR "disabled persons"[All Fields] OR "disabled"[All Fields] OR "disablement"[All Fields] OR "disablements"[All Fields] OR "disabling"[All Fields] OR "disablity"[All Fields])) AND ((ffrft[Filter]) AND (humans[Filter]) AND (1983/1/1:2024/4/12[PDAT]) AND (english[Filter])) | PubMed | 976 |
| SQ12  first-generation professionals | (("rare diseases"[All Fields] OR "rare diseases"[MeSH Terms] OR "rare condition"[All Fields] OR "rare disorder"[All Fields] OR "orphan diseases"[All Fields] OR "neglected diseases"[All Fields]) AND ("first generation"[All Fields] AND ("professional"[All Fields] OR "professional s"[All Fields] OR "professionalism"[MeSH Terms] OR "professionalism"[All Fields] OR "professionality"[All Fields] OR "professionalization"[All Fields] OR "professionalize"[All Fields] OR "professionalized"[All Fields] OR "professionalizing"[All Fields] OR "professionally"[All Fields] OR "professionals"[All Fields]))) AND ((ffrft[Filter]) AND (humans[Filter]) AND (1983/1/1:2024/4/12[PDAT]) AND (english[Filter])) | PubMed | No results found |
| SQ13  first-generation college students | (("rare diseases"[All Fields] OR "rare diseases"[MeSH Terms] OR "rare condition"[All Fields] OR "rare disorder"[All Fields] OR "orphan diseases"[All Fields] OR "neglected diseases"[All Fields]) AND ("first generation"[All Fields] AND ("professional"[All Fields] OR "professional s"[All Fields] OR "professionalism"[MeSH Terms] OR "professionalism"[All Fields] OR "professionality"[All Fields] OR "professionalization"[All Fields] OR "professionalize"[All Fields] OR "professionalized"[All Fields] OR "professionalizing"[All Fields] OR "professionally"[All Fields] OR "professionals"[All Fields]))) AND ((ffrft[Filter]) AND (humans[Filter]) AND (1983/1/1:2024/4/12[PDAT]) AND (english[Filter])) | PubMed | No results found |
| SQ14  employment barriers | (("rare diseases"[All Fields] OR "rare diseases"[MeSH Terms] OR "rare condition"[All Fields] OR "rare disorder"[All Fields] OR "orphan diseases"[All Fields] OR "neglected diseases"[All Fields]) AND (("employability"[All Fields] OR "employable"[All Fields] OR "employer"[All Fields] OR "employer s"[All Fields] OR "employers"[All Fields] OR "employment"[MeSH Terms] OR "employment"[All Fields] OR "employments"[All Fields]) AND ("barrier"[All Fields] OR "barrier s"[All Fields] OR "barriers"[All Fields]))) AND ((ffrft[Filter]) AND (humans[Filter]) AND (1983/1/1:2024/4/12[PDAT]) AND (english[Filter])) | PubMed | 4 |
| SQ15  prison (using prison instead of incarceration as incarceration is also a medical term and yielded results of this condition) | (("rare diseases"[All Fields] OR "rare diseases"[MeSH Terms] OR "rare condition"[All Fields] OR "rare disorder"[All Fields] OR "orphan diseases"[All Fields] OR "neglected diseases"[All Fields]) AND ("prison s"[All Fields] OR "prisoners"[MeSH Terms] OR "prisoners"[All Fields] OR "prisoner"[All Fields] OR "prisons"[MeSH Terms] OR "prisons"[All Fields] OR "prison"[All Fields])) AND ((ffrft[Filter]) AND (humans[Filter]) AND (1983/1/1:2024/4/12[PDAT]) AND (english[Filter])) | PubMed | 8 |
| SQ16  rural areas | (("rare diseases"[All Fields] OR "rare diseases"[MeSH Terms] OR "rare condition"[All Fields] OR "rare disorder"[All Fields] OR "orphan diseases"[All Fields] OR "neglected diseases"[All Fields]) AND (("rural"[All Fields] OR "ruralities"[All Fields] OR "rurality"[All Fields] OR "rurally"[All Fields] OR "ruralness"[All Fields] OR "rurals"[All Fields]) AND ("area s"[All Fields] OR "areas"[All Fields]))) AND ((ffrft[Filter]) AND (humans[Filter]) AND (1983/1/1:2024/4/12[PDAT]) AND (english[Filter])) | PubMed | 65 |
| SQ17  veterans | (("rare diseases"[All Fields] OR "rare diseases"[MeSH Terms] OR "rare condition"[All Fields] OR "rare disorder"[All Fields] OR "orphan diseases"[All Fields] OR "neglected diseases"[All Fields]) AND ("veteran s"[All Fields] OR "veterans"[MeSH Terms] OR "veterans"[All Fields] OR "veteran"[All Fields])) AND ((ffrft[Filter]) AND (humans[Filter]) AND (1983/1/1:2024/4/12[PDAT]) AND (english[Filter])) | PubMed | 84 |
| SQ18  military spouses | (("rare diseases"[All Fields] OR "rare diseases"[MeSH Terms] OR "rare condition"[All Fields] OR "rare disorder"[All Fields] OR "orphan diseases"[All Fields] OR "neglected diseases"[All Fields]) AND (("militaries"[All Fields] OR "military personnel"[MeSH Terms] OR ("military"[All Fields] AND "personnel"[All Fields]) OR "military personnel"[All Fields] OR "military"[All Fields] OR "military s"[All Fields]) AND ("spouse s"[All Fields] OR "spouses"[MeSH Terms] OR "spouses"[All Fields] OR "spouse"[All Fields]))) AND ((ffrft[Filter]) AND (humans[Filter]) AND (1983/1/1:2024/4/12[PDAT]) AND (english[Filter])) | PubMed | No results found |
| SQ19  poverty | (("rare diseases"[All Fields] OR "rare diseases"[MeSH Terms] OR "rare condition"[All Fields] OR "rare disorder"[All Fields] OR "orphan diseases"[All Fields] OR "neglected diseases"[All Fields]) AND ("poverty"[MeSH Terms] OR "poverty"[All Fields] OR "poverty s"[All Fields])) AND ((ffrft[Filter]) AND (humans[Filter]) AND (1983/1/1:2024/4/12[PDAT]) AND (english[Filter])) | PubMed | 237 |
| SQ20  immigrants | (("rare diseases"[All Fields] OR "rare diseases"[MeSH Terms] OR "rare condition"[All Fields] OR "rare disorder"[All Fields] OR "orphan diseases"[All Fields] OR "neglected diseases"[All Fields]) AND ("emigrants and immigrants"[MeSH Terms] OR ("emigrants"[All Fields] AND "immigrants"[All Fields]) OR "emigrants and immigrants"[All Fields] OR "immigrant"[All Fields] OR "immigrants"[All Fields] OR "emigration and immigration"[MeSH Terms] OR ("emigration"[All Fields] AND "immigration"[All Fields]) OR "emigration and immigration"[All Fields] OR "immigration"[All Fields] OR "immigrations"[All Fields] OR "immigrant s"[All Fields] OR "immigrate"[All Fields] OR "immigrated"[All Fields] OR "immigrates"[All Fields] OR "immigrating"[All Fields])) AND ((ffrft[Filter]) AND (humans[Filter]) AND (1983/1/1:2024/4/12[PDAT]) AND (english[Filter])) | PubMed | 36 |
| SQ21  immigrants in USA | (("rare diseases"[All Fields] OR "rare diseases"[MeSH Terms] OR "rare condition"[All Fields] OR "rare disorder"[All Fields] OR "orphan diseases"[All Fields] OR "neglected diseases"[All Fields]) AND (("emigrants and immigrants"[MeSH Terms] OR ("emigrants"[All Fields] AND "immigrants"[All Fields]) OR "emigrants and immigrants"[All Fields] OR "immigrant"[All Fields] OR "immigrants"[All Fields] OR "emigration and immigration"[MeSH Terms] OR ("emigration"[All Fields] AND "immigration"[All Fields]) OR "emigration and immigration"[All Fields] OR "immigration"[All Fields] OR "immigrations"[All Fields] OR "immigrant s"[All Fields] OR "immigrate"[All Fields] OR "immigrated"[All Fields] OR "immigrates"[All Fields] OR "immigrating"[All Fields]) AND "USA"[All Fields])) AND ((ffrft[Filter]) AND (humans[Filter]) AND (1983/1/1:2024/4/12[PDAT]) AND (english[Filter])) | PubMed | 5 |
| SQ22  older age | (("rare diseases"[All Fields] OR "rare diseases"[MeSH Terms] OR "rare condition"[All Fields] OR "rare disorder"[All Fields] OR "orphan diseases"[All Fields] OR "neglected diseases"[All Fields]) AND "older age"[All Fields]) AND ((ffrft[Filter]) AND (humans[Filter]) AND (1983/1/1:2024/4/12[PDAT]) AND (english[Filter])) | PubMed | 56 |
| SQ23  religion | (("rare diseases"[MeSH Terms] OR ("rare"[All Fields] AND "diseases"[All Fields]) OR "rare diseases"[All Fields] OR "rare diseases"[MeSH Terms] OR "rare condition"[All Fields] OR "rare disorder"[All Fields] OR "orphan diseases"[All Fields] OR "neglected diseases"[All Fields]) AND "religion"[All Fields]) AND ((ffrft[Filter]) AND (humans[Filter]) AND (1983/1/1:2024/4/12[PDAT]) AND (english[Filter])) | PubMed | 14 |
| SQ24  Black or African American | (("rare diseases"[All Fields] OR "rare diseases"[MeSH Terms] OR "rare condition"[All Fields] OR "rare disorder"[All Fields] OR "orphan diseases"[All Fields] OR "neglected diseases"[All Fields]) AND "Black or African American"[All Fields]) AND ((ffrft[Filter]) AND (humans[Filter]) AND (1983/1/1:2024/4/12[PDAT]) AND (english[Filter])) | PubMed | 16 |
| SQ26  Asian | (("rare diseases"[All Fields] OR "rare diseases"[MeSH Terms] OR "rare condition"[All Fields] OR "rare disorder"[All Fields] OR "orphan diseases"[All Fields] OR "neglected diseases"[All Fields]) AND "Asian"[All Fields]) AND ((ffrft[Filter]) AND (humans[Filter]) AND (1983/1/1:2024/4/12[PDAT]) AND (english[Filter])) | PubMed | 246 |
| SQ27  Native American Or American Indian | (("rare diseases"[All Fields] OR "rare diseases"[MeSH Terms] OR "rare condition"[All Fields] OR "rare disorder"[All Fields] OR "orphan diseases"[All Fields] OR "neglected diseases"[All Fields]) AND "Native American or American Indian"[All Fields]) AND ((ffrft[Filter]) AND (humans[Filter]) AND (1983/1/1:2024/4/12[PDAT]) AND (english[Filter])) | PubMed | No results found |
| SQ29  Alaska Native | "rare diseases" OR "rare diseases" [MeSH Terms] OR "rare condition" OR "rare disorder" OR "orphan diseases" OR "neglected diseases" AND "Alaska Native" | PubMed | 2 |
| SQ30  Native Hawaiian or Other Pacific Islander | (("rare diseases"[All Fields] OR "rare diseases"[MeSH Terms] OR "rare condition"[All Fields] OR "rare disorder"[All Fields] OR "orphan diseases"[All Fields] OR "neglected diseases"[All Fields]) AND "Alaska Native"[All Fields]) AND ((ffrft[Filter]) AND (humans[Filter]) AND (1983/1/1:2024/4/12[PDAT]) AND (english[Filter])) | PubMed | 3 |
| SQ31  North African persons | (("rare diseases"[MeSH Terms] OR ("rare"[All Fields] AND "diseases"[All Fields]) OR "rare diseases"[All Fields] OR "rare diseases"[MeSH Terms] OR "rare condition"[All Fields] OR "rare disorder"[All Fields] OR "orphan diseases"[All Fields] OR "neglected diseases"[All Fields]) AND "North African persons"[All Fields]) AND ((ffrft[Filter]) AND (humans[Filter]) AND (1983/1/1:2024/4/12[PDAT]) AND (english[Filter])) | PubMed | No |
| SQ32  North African | (("rare diseases"[All Fields] OR "rare diseases"[MeSH Terms] OR "rare condition"[All Fields] OR "rare disorder"[All Fields] OR "orphan diseases"[All Fields] OR "neglected diseases"[All Fields]) AND "North African"[All Fields]) AND ((ffrft[Filter]) AND (humans[Filter]) AND (1983/1/1:2024/4/12[PDAT]) AND (english[Filter])) | PubMed | 7 |
| SQ33  Middle Eastern | (("rare diseases"[All Fields] OR "rare diseases"[MeSH Terms] OR "rare condition"[All Fields] OR "rare disorder"[All Fields] OR "orphan diseases"[All Fields] OR "neglected diseases"[All Fields]) AND "Middle Eastern"[All Fields]) AND ((ffrft[Filter]) AND (humans[Filter]) AND (1983/1/1:2024/4/12[PDAT]) AND (english[Filter])) | PubMed | 7 |
| SQ34  rare diseases AND Hispanics | "rare diseases" OR "rare diseases" [MeSH Terms] OR "rare condition" OR "rare disorder" OR "orphan diseases" OR "neglected diseases" AND "Hispanics" | PubMed | 17 |
| SQ35  race and ethnicity | (("rare diseases"[All Fields] OR "rare diseases"[MeSH Terms] OR "rare condition"[All Fields] OR "rare disorder"[All Fields] OR "orphan diseases"[All Fields]) AND "race"[All Fields] AND "ethnicity"[All Fields]) AND ((ffrft[Filter]) AND (humans[Filter]) AND (1983/1/1:2024/4/12[PDAT]) AND (english[Filter])) | PubMed | 15 |
| SQ36  equity and inclusivity | (("rare diseases"[All Fields] OR "rare diseases"[MeSH Terms] OR "rare condition"[All Fields] OR "rare disorder"[All Fields] OR "orphan diseases"[All Fields]) AND "equity"[All Fields] AND "inclusivity"[All Fields]) AND ((ffrft[Filter]) AND (humans[Filter]) AND (1983/1/1:2024/4/12[PDAT]) AND (english[Filter])) | PubMed | No results |

##### SQ7–36 were derived from the definition of underserved communities Executive Order 13985

##### Abbreviations: MeSH, Medical subject headers; PDAT, Publication Date; SQ, search query.

### Table S2: Keywords used for title screening of publications

| Set | Keywords |
| --- | --- |
| Set 1 | "randomized", "trial", "phase i", "phase ii", "phase iii", "pharmacokinetics", "safety", "efficacy", "effectiveness", "rare disease", "rare disorder", "observational", "systematic", "patient-reported", "outcomes", "survival", "real-world evidence", "dose", "clinical trial", "registry", "registries", "trials", "meta-analysis", "exome", "sequencing", "genomic", "Genomics", "genetics", "genetic", "variants", "congenital" |
| Set 2^a^ | "pregnancy", "pregnant", "Pregnant", "female", "women", "cohort", "Cohort", "population", "cross-sectional", "registry", "registries", "Registry", "caregiver", "Caregiver", "carer", "parent", "parent's", "Parents", "parental", "exome", "sequencing", "genomic", "Genomics", "genetics", "genetic", "disparities", "syndrome", "qualitative", "interview", "congenital", "natal", "prenatal", "pre-natal", "postnatal", "post-natal", "underserved", "religion", "pilgrimage", "poverty", "Veteran", "veterans", "military" |
| Set 3^b^ | Exclusion:“study protocols”, “case reports”, countries other than the US, and non-rare disease terms were also used for exclusion  Inclusion: variations of “registry”, “natural history,” “genetics “genomics”, “sequencing”, “variant”, “mutations” |

##### ^a^Used for results from search queries (SQ7–36; see Table S1) using terms from underserved groups definition.

##### ^b^Manual screening of titles.

### Table S3: Keywords^a^ for identifying RD clinical trials reporting socioeconomic factors data

**Keywords**

"Lesbian", "Gay", "Bisexual", "Transgender", "Queer", "Gender non-conforming", "Non-binary", "Sexual orientation", "Gender identity", "Parent", "Caregiver", "Marital status", "Married", "Divorced", "Partner", "Single", "Widowed", "Employment", "Financial status", "Working status", "Income", "Poverty", "Occupation", "Education", "Language", "English proficiency", "Spanish", "Primary Language", "Residence type", "Rural", "Urban", "Residence", "Housing type", "Disability", "Disable", "Disabled", "Disabilities", "Insurance", "Pregnancy", "Pregnant", "Veteran", "Prisoner", "Incarceration", "Jail", "Military", "Immigrant", "Religion", "Belief system"

##### ^a^Lexical variants were created to include plural forms. Abbreviations: RD, rare disease.

### Table S4: A snapshot^a^ of US-based RD publications reporting race and ethnicity in a non-standard format

| **PMID/PMCID** | **Title** | **Year** | **Details** |
| --- | --- | --- | --- |
| PMC4030757 | Efficacy and safety of rifampicin for multiple system atrophy: a randomised, double-blind, placebo-controlled trial | 2014 | Uses White and non-White as race subcategories |
| PMC6500220 | Prevalence and practice for rare diseases in primary care: a national cross-sectional study in the USA | 2019 | Race: Non-Hispanic Black, Non-Hispanic White, Hispanics, Others |
| 34096130 | “Doctors can read about it, they can know about it, but they’ve never lived with it”: How parents use social media throughout the diagnostic odyssey | 2021 | Uses the term Caucasian as a race sub category |
| 34688299 | Development of therapies for rare genetic disorders of GPX4: roadmap and opportunities | 2021 | Race: Iranian, Caucasian, Black, Pakistani, Indian, Middle Eastern, Yemeni, Turkish, Armenian, Iraqi, Indian |
| - PMC9892172 | Rare diseases, common barriers: disparities in pediatric clinical genetics outcomes | 2023 | Ancestry: African continent, Asia/Pacific Islands, Middle East, Latin America, North America, Europe and Unknown |
| 35653343 | Rare diseases, common barriers: disparities in pediatric clinical genetics outcomes | 2023 | Combined the Asian and Native Hawaiian or Other Pacific Islander race subcategories |
| 36949506 | Sources of variation in estimates of Duchenne and Becker muscular dystrophy prevalence in the United States | 2023 | Race/Ethnicity: Black, White, Other, and Hispanic. Other included race other than Black, Hispanic, or White, including multiple races and missing |
| PMC10620944 | Biallelic PRMT7 pathogenic variants are associated with a recognizable syndromic neurodevelopmental disorder with short stature, obesity, and craniofacial and digital abnormalities | 2023 | Uses country of origin as ethnicity |

##### ^a^non-exhaustive list.

##### Abbreviations: PMCID, PubMed Central Identifier; PMID, PubMed Identifier; RD, rare disease; US, United States.

### Table S5: Socioeconomic factors reporting in US-based RD publications

|  | **US-only publications** | | **Multinational^a^ publications** | |
| --- | --- | --- | --- | --- |
| **Variable** | **Frequency or**  **number of publications** | **Range of participant proportions** | **Frequency or**  **number of publications** | **Range of participant proportions** |
| **Socioeconomic factors** | 21 | NA | 9 | NA |
| **Language** | 9 | NA | 7 | NA |
| English | 9 | 66.7–100% | 4 | 93–100% |
| Spanish | 4 | 3.1–33.3% | 0 |  |
| French | 0 | NA | 2 | French=7% (Canadian and  Belgian dialects) |
| German | 0 | NA | 2 | NA |
| Other Languages | 3 | Kurdish=0.3%,  Arabic=4.7%,  Others=3.7% | 2 | Italian, Dutch  (Netherlands and Flemish dialects) |
| **Education** | 16 |  |  |  |
| Basic education | 10 | 0–62% | 3 | 1– 41.2% |
| Some college or Associate degree | 10 | 10–33% | 5 | 17.7–42.4% |
| College graduate | 9 | 23.3–87.1% | 3 | 28.5–60.3% |
| Advanced education | 12 | 7–58.3% | 5 | 14– 28.4% |
| Education in years | 0 | NA | 2 | 15.2–15.8 years (mean) |
| **Income** | 12 | NA | 2 | NA |
| Below federal poverty level (<$30K) | 8 | 7.8–85.2% | 2 | 4–8% |
| >200% of poverty level | 1 | 4.5% | NA | NA |
| Lower-middle income ($30–$50K) | 6 | 7–26% | 2 | 10.7–16.1% |
| Upper-middle income ($50–$100K) | 7 | 11–48.6% | 2 | 9.6–23.9% |
| High income ($100–$150K) | 6 | 14–30.8% | 2 | 36–36.8% |
| Very high income (>$150K) | 5 | 7–23.3% | NA | NA |
| Unrecorded/Missing/Not asked/  Don’t know | 4 | 1–8% | 2 | 4.7–10.8% |
| **Employment** | 9 | NA | 4 | NA |
| Full-time Employment | 9 | 14–73.3% | 2 | 28.1–43.9% |
| Part-time Employment | 6 | 7–42.2% | 1 | 24.4% |
| Unemployed/Disabled | 6 | 9.7–36.7% | 1 | 37.2–55.8% |
| Other employment status or Employment skill level | 4 | 1–29% | 1 | 3.4–66.7% |
| Missing data | 2 | 1– 2.8% | 0 | NA |
| **Geographical location** | 6 | NA | 1 | NA |
| Urban | 3 | 15–84.5% | 1 | 56.1% |
| Rural | 3 | 1.6–15.5% | NA | NA |
| Suburban or Rural | NA | NA | 1 | 43.9% |
| Suburban | 1 | 19% | NA | NA |
| Metro | 1 | 80% | NA | NA |
| Not reported/unrecorded | 2 | 1.6–3% | NA | NA |
| Distance traveled to clinic | 1 | <50 miles=41%  50–99 miles=24%  100–249 miles=15%  ≥250 miles=16% | NA | NA |
| Average distance from research site | 1 | 42–320 miles | NA | NA |
| Geographical location in the US | 1 | Northeast=19.9%  Midwest=15.8%  Southwest=7.0%  West=23.8%  Other/Unknown=17.0% | NA | NA |
| **Insurance** | 12 | NA | 2 | NA |
| Private/Commercial Insurance | 10 | 3.8–100% | NA | NA |
| Public Insurance (Medicaid/Medicare/Government Assistance) | 12 | 2.7–67.6% | NA | NA |
| Military insurance | 1 | 13.10% | NA | NA |
| Self-Pay/No insurance | 4 | 1–6.2% | NA | NA |
| Other/Unknown insurance | 5 | 0.2–6.6% | NA | NA |
| Insurance coverage | 1 | 98.8% | 2 | 92.5–93.5% |
| **Relationship status** | 5 | NA | 4 | NA |
| Married/Committed | 5 | 55.8–82.1% | 3 | 67.6–81.5% |
| Single/Not in a relationship | 2 | 18–21% | 1 | 32.4% |
| Divorced | 1 | 5% | NA | NA |
| Other | 1 | 17.9% | NA | NA |
| **Social Support** |  |  |  |  |
| Special education assistance | 1 | 71% | NA | NA |
| Food assistance | 1 | 16% | NA | NA |
| **Disability allowance** | 3 | 27–100% | NA | NA |

##### ^a^Publications reporting studies conducted in the US and other countries.

##### Abbreviations: NA, not applicable; RD, rare disease; US, United States. Table S6: Keyword–condition matches of RD clinical trials

| **Similarity Range (%)** | **Non-acronyms NCTs, N (%)** | **Acronym NCTs, N (%)** | **Total NCTs, N (%)** |
| --- | --- | --- | --- |
| 100 | 6223 (77.4) | 50 (11.4) | 6273 (74.0) |
| >80–100 | 795 (9.0) | 8 (1.8) | 803 (9.5) |
| >60–80 | 649 (8.1) | 2 (0.5) | 651(7.7) |
| >40–60 | 244 (3.0) | 11(2.5) | 255 (3.0) |
| >20–40 | 45 (0.6) | 216 (49.2) | 261 (3.1) |
| 0–20 | 0 (0.0) | 132 (30.1) | 132 (1.6) |
| Non-specific acronym matches | - | 369 (4.3)^a^ | - |
| Total NCTs | 8036 (100.0) | 439 (100.0) | 8475 (100.0) |

##### ^a^Aggregate of keyword–condition matches with similarity score <100%.

##### ^b^Percentage calculated for the total number of NCTs.

##### Abbreviations: N number of trials; RD, rare disease; NCT, National Clinical Trial Identifier.

### Table S7: Participant distribution statistics from US-based RD clinical trials (1986–2022)^a^

| **Variable** | **Total, n (%)** | **US-only trials, n (%)** | **Multinational^a^ trials, n (%)** |
| --- | --- | --- | --- |
| **Age^b^** | | | |
| Total age | 1487552 (100.0) | 889317 (100.0) | 599435 (100.0) |
| Child | 131365 (8.8) | 46816 (5.3) | 84549 (14.1) |
| Adult | 40196 (2.7) | 25760 (2.9) | 14648 (2.4) |
| Older adult | 9284 (0.6) | 4960 (0.6) | 4324 (0.7) |
| Child and Adult | 95754 (6.4) | 35953 (4.1) | 59831 (10.0) |
| Adult and Older Adult | 664983 (44.7) | 310035 (34.9) | 355566 (59.3) |
| Child, Adult and Older adult | 545970 (36.7) | 465793 (52.4) | 80517 (13.4) |
| **Sex or Gender^c^** | | | |
| Total Sex or Gender | 1447552 (100.0) | 860934 (100.0) | 586618 (100.0) |
| Male | 665896 (46.0) | 362289 (42.1) | 303607 (51.8) |
| Female | 781635 (54.0) | 498624 (57.9) | 283011 (48.2) |
| Other or unknown | 21 (0.001) | 21 (0.002) | 0 (0.0) |
| **Race (NIH/OMB)^d^** | | | |
| Total race | 401178 (100.0) | 162202 (100.0) | 238976 (100.0) |
| American Indian or Alaska Native | 2728 (0.7) | 850 (0.5) | 1878 (0.8) |
| Asian | 30032 (7.5) | 5125 (3.2) | 24907 (10.4) |
| Black or African American | 45793 (11.3) | 23707 (14.6) | 22086 (9.2) |
| Native Hawaiian or Other Pacific Islander | 1019 (0.3) | 453 (0.3) | 566 (0.2) |
| White | 295163 (73.6) | 123284 (74.4) | 171879 (71.8) |
| More than one race | 6173 (1.5) | 1888 (1.2) | 4285 (1.2) |
| Race unknown or not reported | 20270 (5.1) | 6895 (4.3) | 13375 (5.6) |
| **Ethnicity (NIH/OMB)^d^** | | | |
| Total ethnicity | 353800 (100.0) | 134585 (100.0) | 219215 (100.0) |
| Hispanic or Latino | 38769 (11.0) | 12258 (9.1) | 26511 (12.1) |
| Not Hispanic or Latino | 290218 (82.0) | 115426 (85.8) | 174792 (79.7) |
| Ethnicity unknown or not reported | 24813 (7.0) | 6901 (5.1) | 17912 (8.2) |

##### ^a^Trials conducted in the US and other countries.

^b^Excluding trials with ‘complex data’

##### ^c^Excluding trials reporting sex or gender in a customized format and trials with complex data.

##### ^d^Trials reporting race and ethnicity in NIH/OMB format and trials with complex data.

##### Abbreviations: OMB, Office of Management and Budget; n, number of participants; NIH, National Institute of Health; RD, rare disease; US, United States.

### Table S8: Socioeconomic factors reporting in US-based RD trials (1991–2022)^a^

| **Socioeconomic factors** | **Number and percentage of trials, n (%)** |
| --- | --- |
| Education | 8 (50.0) |
| Employment/Occupation | 4 (25.0) |
| Insurance | 6 (37.5) |
| Income | 4 (25.0) |
| Relationship status | 11 (68.8) |
| Religion | 2 (12.5) |

##### ^a^Refers to trial start years.

### Table S9: Participant distribution statistics in US-based RD trials reporting socioeconomic factors (1991–2022)^a^

|  | **US-only trials** | | **Multinational^b^ trials** | |
| --- | --- | --- | --- | --- |
|  | **Number of trials** | **Range of participant proportions** | **Number of trials** | **Range of participant proportions** |
| **Socioeconomic factors** | 14 | NA | 2 | NA |
| **Education** |  |  |  |  |
| Primary school | 5 | 0.7– 60% | 0 | 0 |
| High school | 8 | 15.8–41.1% | 0 | 0 |
| Some college | 7 | 27.2–60.1% | 0 | 0 |
| Degree | 5 | 18.2–66.7% | 0 | 0 |
| Masters | 1 | 13.3% | 0 | 0 |
| Doctoral | 1 | 1.4% | 0 | 0 |
| **Employment/Occupation** |  |  |  |  |
| Employed | 3 | 32.4–63.8% | 0 | 0 |
| Not employed | 3 | 10.8–64.8% | 0 | 0 |
| Retired | 1 | 10.40% | 0 | 0 |
| Homemaker | 1 | 5% | 0 | 0 |
| Agriculture | 0 | 0 | 1 | 37.1% |
| Non-agriculture | 0 | 0 | 1 | 55.7% |
| Disabled | 2 | 5–20% | 0 | 0 |
| **Insurance** |  |  |  |  |
| Private/Commercial Insurance | 6 | 22– 83.8% | 0 | 0 |
| Public Insurance (Medicaid/Medicare) | 5 | 7.5– 59.4% | 0 | 0 |
| Government-subsidized insurance | 1 | 76.3% | 0 | 0 |
| Self-paid/none | 4 | 1.7– 3.6% | 0 | 0 |
| **Income** |  |  |  |  |
| <$10K | 1 | 6.10% | 0 | 0 |
| $10–$20K | 1 | 9.3% | 0 | 0 |
| $20–$35K | 1 | 11.1% | 0 | 0 |
| $35–$50K | 1 | 9.7% | 0 | 0 |
| $50–$75K | 1 | 20.4% | 0 | 0 |
| >$75K | 2 | 4.3– 50% | 0 | 0 |
| **Relationship Status** |  |  |  |  |
| Married/ living with partner | 11 | 4.3– 83.6% | 0 | 0 |
| Single | 10 | 1.3– 82.6% | 0 | 0 |
| Divorced/Separated/ | 8 | 1.3– 34.8% | 0 | 0 |
| Widowed |  |  |  |  |
| **Religion** |  |  |  |  |
| Jewish (Ashkenazi) | 0 | 0 | 1 | 45.2% |
| Non-Jewish | 0 | 0 | 1 | 48.4% |

##### ^a^Refers to trial start years.

##### ^b^Trials conducted in the US and other countries.

##### Abbreviations: n, number of trials; NA, not applicable; RD, rare disease; US, United States.

### Table S10: p-values for statistical significance of aggregate and mean race and ethnicity proportions, in US-only RD trials (1991–2022)^a^, compared with the US census averages^b^

|  | **Aggregate proportions (p-values)** | **Mean proportions (p-values)** |
| --- | --- | --- |
| **Race** | | |
| American Indian or Alaska Native | 1.9408e-90* | 3.3896e-139* |
| Asian | 7.4601e-217* | 1.5631e-100* |
| Native Hawaiian or Other Pacific Islander | 4.1747e-16* | 4.1080e-01 |
| Black or African American | 3.1111e-126* | 2.8697e-30* |
| White | 1.7737e-214* | 0.0000e+00* |
| More than one race | 0.0000e+00* | 0.0000e+00* |
| Race unknown or not reported | 3.2089e-25* | 5.5431e-02 |
| **Ethnicity** | | |
| Hispanic or Latino | 0.0000e+00 | 0.0000e+00* |
| Not Hispanic or Latino | 1.9985e-37 | 4.0347e-74* |
| Unknown or not reported | Skipped due to zero count in the census | Skipped due to zero count in the census |

##### ^a^Refers to trial start years.

##### ^b^Weighted averages were calculated for 1990–2022 based on the participant distribution in the rare disease clinical trials.

##### *Indicates significance at Bonferroni-corrected significance level (race: 0.0071; ethnicity: 0.0167)

##### Abbreviations: RD, rare disease; US, United States.

### Table S11: p-values for statistical significance of aggregate and mean proportions of race and ethnicity in US-only RD trials (1991–2022)^a^, compared with US census averages^b^: by study phase and funder type

|  | **Race: aggregate proportions (p-values)** | **Ethnicity: aggregate proportions (p-values)^c^** | **Race: mean proportions**  **(p-values)** | **Ethnicity: mean proportions (p-values)^c^** |
| --- | --- | --- | --- | --- |
| **Study phase vs Census** | | | | |
| Early Phase1 vs Census | 5.43e-03* | 0.00e+00* | 2.9751e-03* | 0.0000e+00* |
| Not applicable vs Census: | 0.00e+00* | 0.00e+00* | 7.3109e-126* | 0.0000e+00* |
| Phase1 vs Census | 1.48e-30* | 0.00e+00* | 3.7341e-35* | 0.0000e+00* |
| Phase1\|Phase2 | 3.07e-152* | 0.00e+00* | 1.1197e-114* | 0.0000e+00* |
| Phase2 vs Census | 0.00e+00* | 0.00e+00* | 0.0000e+00* | 0.0000e+00* |
| Phase2\|Phase3 | 3.26e-32* | 0.00e+00* | 1.1537e-30* | 0.0000e+00* |
| Phase3 vs Census: | 0.00e+00* | 0.00e+00* | 4.2891e-270* | 0.0000e+00* |
| Phase4 vs Census | 2.41e-229* | 0.00e+00* | 7.3669e-48* | 0.0000e+00* |
| **Funder type vs Census** | | | | |
| Federal vs Census | 8.85e-17* | 0.00e+00* | 2.1359e-26* | 0.00e+00* |
| Industry vs Census | 0.00e+00* | 0.00e+00* | 6.7123e-272* | 0.00e+00* |
| NIH vs Census | 2.46e-209* | 0.00e+00* | 1.2794e-146* | 0.00e+00* |
| Other vs Census** | 0.00e+00 | 0.00e+00* | 0.0000e+00* | 0.00e+00* |

##### ^a^Refers to trial start years.

##### ^b^Weighted averages were calculated for the years 1990–2022 based on the participant distribution in the rare disease clinical trials;

##### ^c^All likely zero because the US census does not have an unknown or not reported category.

##### *Indicates significance at 0.05.

##### Abbreviations: NIH, National Institute of Health, RD, rare disease; US, United States
